# Impact of Non-Contrast Enhanced Imaging Input Sequences on the Generation of Virtual Contrast-Enhanced Breast MRI Scans using Neural Networks

**DOI:** 10.1101/2024.05.03.24306067

**Authors:** Andrzej Liebert, Hannes Schreiter, Lorenz A Kapsner, Jessica Eberle, Chris Ehring, Dominique Hadler, Luise Brock, Ramona Erber, Julius Emons, Frederik B. Laun, Michael Uder, Evelyn Wenkel, Sabine Ohlmeyer, Sebastian Bickelhaupt

**Affiliations:** Institute of Radiology, Universitätsklinikum Erlangen, Friedrich-Alexander-Universität Erlangen-Nürnberg (FAU), Erlangen, Germany; Lehrstuhl für Medizinische Informatik, Friedrich-Alexander-Universität Erlangen-Nürnberg (FAU), Germany; Institute of Pathology, Universitätsklinikum Erlangen, Erlangen, Comprehensive Cancer Center Erlangen-EMN, Friedrich-Alexander-Universität Erlangen-Nürnberg (FAU), Germany; Department of Gynecology and Obstetrics, Erlangen University Hospital, Comprehensive Cancer Center Erlangen-EMN, Friedrich-Alexander-Universität Erlangen-Nürnberg (FAU), Erlangen, Germany; Medizinische Fakultät, Friedrich-Alexander-Universität Erlangen-Nürnberg (FAU), Erlangen, Germany. Radiologie München, München, Germany; German Cancer Research Center (DKFZ), Heidelberg, Germany

## Abstract

**Background:** Virtual contrast-enhanced (vCE) imaging techniques are an emerging topic of research in breast MRI.

**Purpose:** To investigate how different combinations of T1-weighted (T1w), T2-weighted (T2w), and diffusion-weighted imaging (DWI) impact the performance of vCE breast MRI.

**Materials and Methods:** The IRB-approved, retrospective study included 1064 multiparametric breast MRI scans (age:52±12 years) obtained from 2017-2020 (single site, two 3T MRI). Eleven independent neural networks were trained to derive vCE images from varying input combinations of T1w, T2w, and multi-b-value DWI sequences (b-value=50–1500s/mm^2^). Three readers evaluated the vCE images with regards to qualitative scores of diagnostic image quality, image sharpness, satisfaction with contrast/signal-to-noise-ratio, and lesion/non-mass enhancement conspicuity. Quantitative metrics (SSIM, PSNR, NRMSE, and median symmetrical accuracy) were analyzed and statistically compared between the input combinations for the full breast volume and both enhancing and non-enhancing target findings.

**Results:** The independent test set consisted of 187 cases. The quantitative metrics significantly improved in target findings when multi-b-value DWI sequences were included during vCE training (p<.05). Non-significant effects (p>.05) were observed for the quantitative metrics on the full breast volume when comparing input combinations including T1w. Using T1w and DWI acquisitions during vCE training is necessary to achieve high satisfaction with contrast/SNR and good conspicuity of the enhancing findings. The input combination of T1w, T2w, and DWI sequences with three b-values showed the best qualitative performance.

**Conclusion:** vCE breast MRI performance is significantly influenced by input sequences. Quantitative metrics and visual quality of vCE images significantly benefit when a multi b-value DWI is added to morphologic T1w-/T2w-sequences as input for model training.

**Key Results:**

1. The inclusion of diffusion-weighted imaging significantly improves the conspicuity of lesions/non-mass enhancements and satisfaction with the image contrast in virtual contrast-enhanced breast MRI.
2. The quality of virtual contrast-enhanced breast MRI scans benefits from the inclusion of high-resolution morphologic T1-weighted image acquisitions.
3. Quantitative metrics such as the structural similarity index and peak signal-to-noise ratio calculated over the entire breast volume insufficiently reflect variations in lesion/non-mass enhancement’s individual characteristics.

## INTRODUCTION

Breast MRI routinely includes the acquisition of contrast-enhanced (CE) image series (1) after intravenous injection of gadolinium-based contrast agents (GBCAs). GBCAs help contrast tissue changes associated with altered neoangiogenesis and/or extracellular GBCA distribution patterns frequently found in significant findings. However, the necessity of GBCA administration might limit accessibility, especially in screening settings (2–5). GBCA administration increases the (direct/indirect) costs of breast MRI, challenging the cost-effectiveness during screening in low- and moderate-risk populations (2,6,7), since its additive time for patient preparation is inevitable even if MRI scanning is shortened (e.g., in abbreviated approaches) (8,9) and the costs of the GBCA itself retained (10–12). Furthermore, gadolinium depositions in the human body have been described in literature (13) potentially constituting a barrier to annual screenings.

The generation of virtual contrast-enhanced (vCE) MRI scans from unenhanced acquisitions using deep learning was first reported in brain studies (14–19). However, several recent publications (20–26) have shown the technical feasibility of such approach for breast imaging. Given the novelty of this research field, it is expected that a high variability is yet observed in the literature with regards to the choice of input data used to train vCE imaging neural networks. T1-weighted (T1w) sequences (20,21,23) have been used for vCE breast MRI as well as combinations of T1w and T2-weighted (T2w) image acquisitions (22) or more complex protocols including diffusion-weighted imaging (DWI)(26).

Our study aims at systematically investigating the impact of different MRI input sequences on the ability of neural networks to generate vCE breast MRI scans using quantitative global and regional metrics in both enhancing and non-enhancing findings. A multi-reader study was also conducted to investigate the image quality and lesion/non-mass enhancement (NME) conspicuity.

## MATERIALS AND METHODS

### Summary of Patient Cohort, MRI Protocol, and Semantic Enrichment

This retrospective study was IRB approved and the need for informed consent was waived. The study included 1064 clinically indicated breast MRI scans (mean age 52 years, standard deviation 12 years) acquired from 2017-2020 at a single site. The examinations were performed using two clinical routine 3T MRI scanners (MAGNETOM Skyra-Fit or MAGNETOM Vida, Siemens Healthineers) in the prone position using a dedicated 18-channel breast coil (Siemens Healthineers). The examination protocol included T1w, T2w, multi-b-value DWI (b-values: 50, 750, 1500 s/mm^2^), and five-timepoint DCE image acquisitions consisting of T1w image acquisition before and five acquisitions after intravenous GBCA administration (gadobutrol; Bayer, Leverkusen, Germany; 0.1 mmol/kg-body weight, injection speed=2 mL/s). Subtraction series were automatically derived by subtracting the image of the native T1w image acquisition from the DCE T1w image acquisitions. Sequence details are given in the Table 1.

Ninety-one MRI scans were excluded owing to the presence of breast implants (Figure 1a). The final cohort (n=973) was randomly divided at the patient level into training (n=710), validation (n=76), and separate independent test sets (n=187).

The independent test set was evaluated in two experiments (Figure 1b).

- <u>Experiment 1: Quantitative analyses</u> were performed for the full test dataset for a) the entire-breast volume (n=187) and b) using a subset of the examinations in which target findings, either enhancing or non-enhancing, could be identified and subsequently manually segmented (145/187 cases), as described in the Quantitative Analysis subsection.
- <u>Experiment 2: Qualitative analyses</u> were performed via visual readings of three independent readers using the full independent test set (n=187) and corresponding scores described in the Qualitative Analysis subsection.

All cohort examinations (n=973) were previously included in studies focused on detecting artifacts in dynamic contrast-enhanced and DWI derived maximum-intensity projections (27,28).

**Figure 1a:**
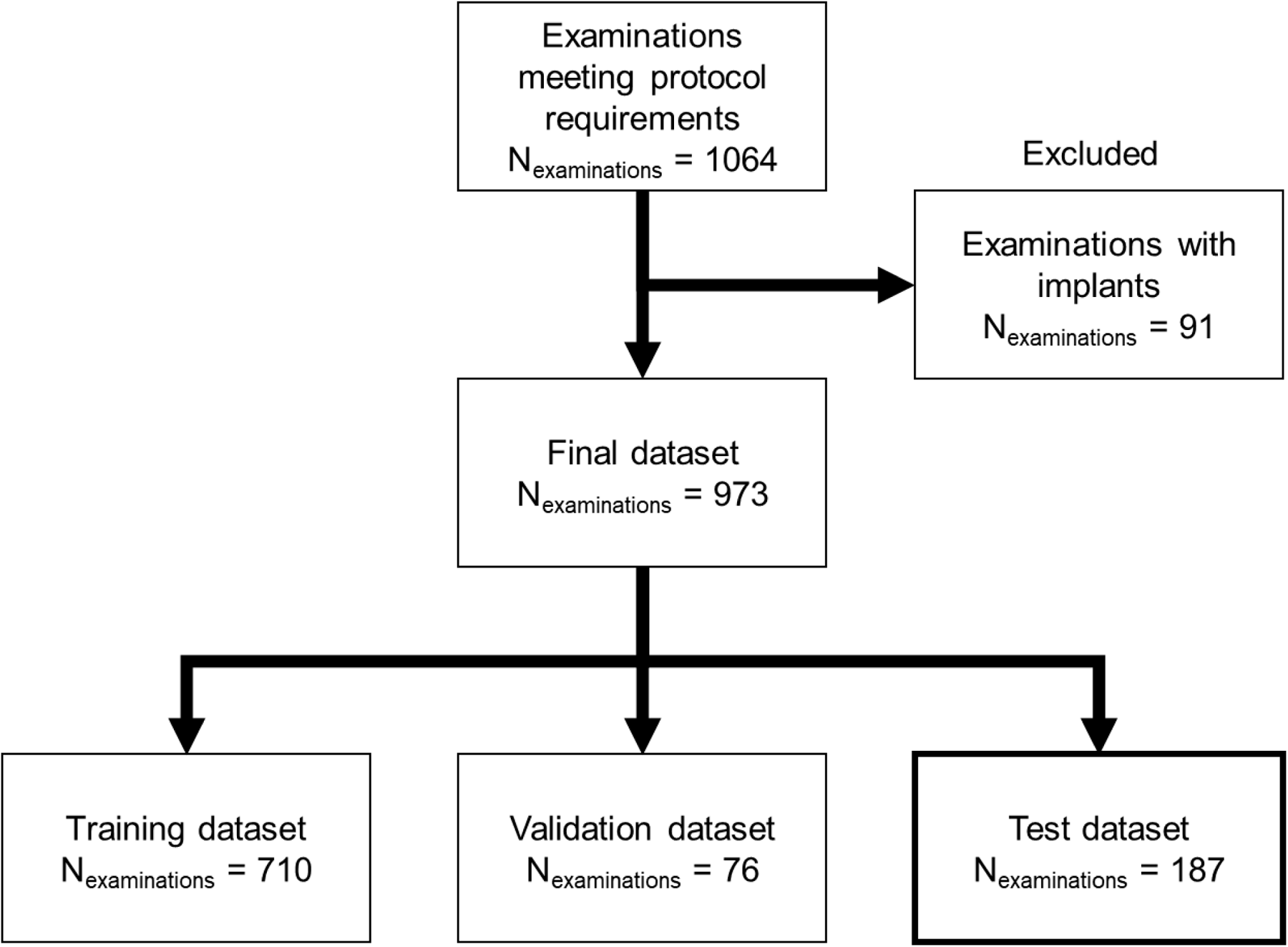
The full cohort consisted of all patients who underwent diagnostic breast MRI on one of two clinical 3T scanners (MAGNETOM Skyra-Fit or MAGNETOM Vida, Siemens Healthineers) from January 2017 to June 2022. The protocol included a five-point dynamic contrast enhanced acquisition, a T1-weighted image acquisition, a T2-weighted fat-saturated image acquisition, and a multi-b-value diffusion-weighted image acquisition containing b-values of 50, 750, and 1500 s/mm^2^.

**Figure 1b:**
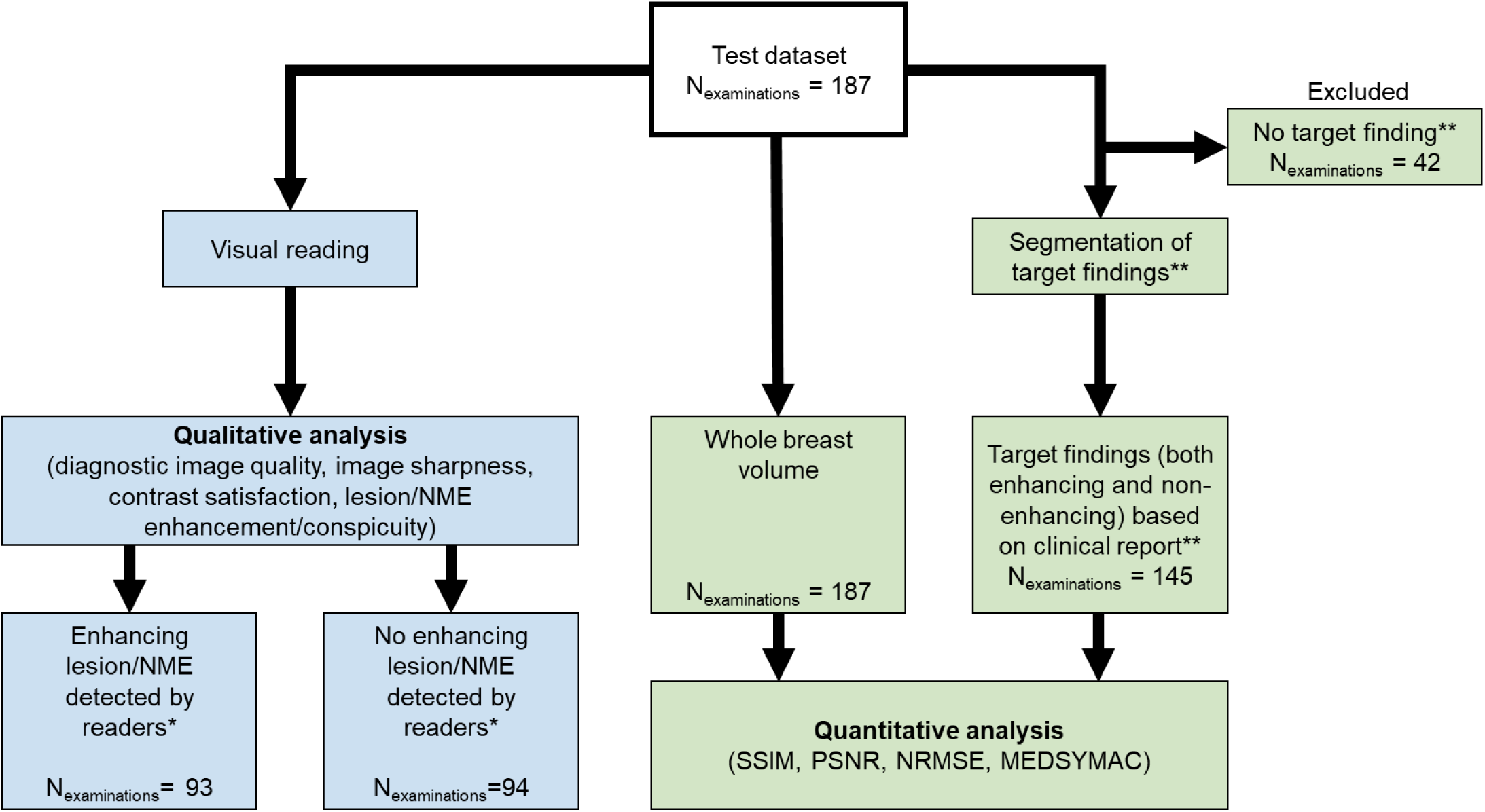
Flow chart of the two experiments conducted. Visual reading was performed by three independent readers. *Enhancing lesions/NMEs were stratified according to the majority vote of the three readers. **Target findings refer to the findings within the examination that could be both benign or malignant and non-mass or mass enhancement as well as findings not enhancing (e.g., cysts) but morphologically delineated from healthy fibro-glandular tissue

### Literature Review of Neural Network Approaches and Input Sequences Used for vCE Breast MRI

A literature review was conducted including peer-reviewed articles/conference abstracts to establish the current knowledge on vCE breast MRI. The primary focus was to identify MRI sequences utilized as input data, specific neural network architectures implemented, and experiments performed for both quantitative and qualitative evaluations of results. The search methodology is detailed in the Supplement Material section “Literature Review”. The literature review is summarized in Table 2. The most commonly used MRI input sequence for generating vCE MRI scans was a morphologic unenhanced T1w. Half of the studies had also reported incorporating morphologic T2w sequences; further two used DWI sequences, and one used apparent diffusion coefficient (ADC) maps.

As our breast MRI dataset contained all three types of native input acquisitions (T1w, T2w, and DWI) described in the literature, we investigated the use of all possible different combinations of these inputs. As our dataset included advanced DWI acquisitions with b-values of 50, 750, and 1500 s/mm^2^ (further called b50, b750, and b1500, respectively), the b-values were grouped as input channels of b50/b750 and b50/b750/b1500, thus reflecting clinically used combinations. The rationale for excluding the ADC maps and simulated low-dose images is shown in the Supplement Material section “Rationale for the Exclusion of ADC and Simulated Low-Dose Images”. In order to investigate the impact of the individual input combinations we decided to investigate all possible combinations of the T1w, T2w and the two DWI input sets. This resulted in the following combinations of non-contrast-enhanced input sequences:

1. T1w
2. T1w, T2w
3. T1w, b50/b750
4. T1w, b50/b750/b1500
5. T2w
6. T2w, b50/b750
7. T2w, b50/b750/b1500
8. b50/b750
9. b50/b750/b1500
10. T1w, T2w, b50/b750
11. T1w, T2w, b50/b750/b1500

The combinations including just T2w, just DWI sequences or the combinations of these two were not previously proposed in other works. We decided to include these into the evaluation due to their potential application in non-contrast enhanced abbreviated protocols (29,30).

### Neural Network Architecture and Training

The two predominant architectures used in generating vCE breast MRI scans were generative adversarial networks (GANs) and encoder–decoder convolutional neural networks (Table 1). As this work sought to investigate the impact of MRI input sequences on the ability of neural networks to generate accurate and clinically relevant vCE MRI scans, we employed a 2D-U-net architecture with three encoder and decoder stages, an established encoder–decoder network, for our analysis.

The rationale for selecting the U-net architecture was based on its robustness compared with GAN-based solutions. By employing a simpler and more robust architecture, we aimed to minimize potential confounding factors introduced by more complex models such as the necessity to optimize discriminator networks in GANs.

During training, single slices of the respective combinations of the pre-contrast MRI acquisitions were used as input channels of the neural network. The second post-contrast subtraction of the dynamic contrast-enhanced series was selected as the ground truth. The 2D-U-Net architecture implementation, data preprocessing, and network training are detailed in the Supplement Figure 1 and Supplement Material section “Neural Network Architecture and Training”.

### Quantitative Analysis

The independent test set was further enriched for quantitative evaluations through adding manual segmentations as laid out in detail in the Supplemental Material section “Segmentation of Target Findings”. In short, the segmentation was performed by a medical student with 2 years of experience in breast MRI research under the supervision of a board-certified radiologist (D.H.>10 years of experience), using the open-source 3D Slicer Software’s [version 4.11] (31) built-in region draw function. During this segmentation the student was blinded to the results of the 11 vCE neworks. The choice of target findings for segmentation was based on the radiological reports generated during clinical routine reading. Such findings included both clinically significant and insignificant findings independent of the GBCA-uptake (e.g., cysts, fibroadenomas, hematomas, intramammary lymph nodes, atheromas, lipoid necroses, and focal areas of mastopathy).

For quantitative evaluation of the image series, two similarity metrics [structural similarity index (SSIM) (32) and– peak signal-to-noise ratio (PSNR)] and two error metrics [normalized root mean square error (NRMSE) and median symmetrical accuracy (MEDSYMAC) (33)] were calculated. All metrics were evaluated within the full breast volume and in the individual segmentations created on both enhancing and non-enhancing target findings. The target finding segmentations and choice of the metrics are detailed in the Supplement Material section “Quantitative Metrics Choice and Calculation”.

### Qualitative Analysis

The literature review allowed the definition of representative qualitative evaluations. The most common qualitative evaluation was a side-by-side comparison of CE and vCE images. The qualitative features evaluated were the diagnostic image quality, image sharpness, satisfaction with the image to fulfill its diagnostic task (allowing for visual depiction of lesions/NMEs and visual signal-to-noise ratio), and lesion/NME conspicuity of the vCE images compared with the CE images.

The reading tasks were as follows: The vCE and CE images were evaluated side by side using the single slices from the original CE image acquisition and the corresponding slice from all 11 vCE input combination variations. All evaluations were performed by three independent readers (two board-certified radiologists S.B. and D.H. with >10 years of experience each and one medical student J.E. with 2 years of experience in breast MRI research) using multi-point Likert-like scales. The readers were not informed about the specific input sequences used to generate each evaluated contrast. However, due to their expertise in breast MRI, the readers might have inferred some of the likely input combinations, since for instance, vCE images created solely from DWI inputs retained a noticeable “DWI-like” texture and e.g. certain subtle yet discernible textural features in the CE images made them distinguishable from the vCE images (minor non-diagnostically significant but visible) artifacts that are typically present in CE subtractions and a more blurred appearance of background parenchymal enhancement in the vCE images). During reading, the following features were evaluated for both CE and vCE images using an 11-point Likert-like scale (0 = non-acceptable/insufficient, 10 = excellent): diagnostic image quality, image sharpness, and satisfaction with image contrast and visual signal-to-noise ratio.

Further, potentially significant enhancing lesions or NMEs were compared with the surrounding tissue and evaluated using a 9-point Likert-like scale for lesion/NME conspicuity (0= lesion/NME not visible, 8 = perfect lesion/NME conspicuity) on the CE subtraction images. For the vCE images, the readers were asked whether the enhancing lesion/NME was correspondingly reflected regarding its conspicuity using an 11-point Likert-like scale (0 = lesion/NME not visible, 8 = fully identical lesion/NME conspicuity compared with GBCA subtraction; expanded by the two scores of 9 = lesion/NME enhancing stronger than on CE subtraction images and 10=lesion/NME enhancing much stronger than on CE images). The more extensive Likert-like scale was selected to allow for a more granular assessment of the data, enabling more detailed evaluations.

### Statistical Analysis

Differences in the ordered variables between the generated vCE images and CE ground truth images were evaluated using the Friedmann’s test, followed by a post-hoc Nemanyi test in significant cases. The resulting p-values were adjusted for multiple comparisons using the Bonferroni method and a p-value of 0.05 was considered as significant. Differences in the quantitative scores were determined both for metrics in the entire breast and segmented findings. Differences in the qualitative scores were evaluated both for all patients and patients among whom an enhancing lesion/NME was identified in the original CE image after a majority voting by all three readers. All statistical analyses were performed using Python (version 3.9.13).

**Table 1:** MRI Protocol.

Impact of Imaging Input Sequences on the Generation vCE Breast MRI
| Table 1: MRI Protocol |  |  |  |  |  |  |  |  |  |  |
| --- | --- | --- | --- | --- | --- | --- | --- | --- | --- | --- |
| Sequence | Sequence Type | Matrix Size | FoV (mm×mm) | Slice thickness (mm) | TR (ms) | TE (ms) | IR (ms) | FA (°) | No. of Averages | Fat Saturation |
| T1w* | 3D-GRE | 448×448×112–128 | 360×360–430×430 | 1.5–1.8 | 5.97 | 2.46 | - | 10 | 1 | None |
| T2w | 2D-SE | 448×448×34–49 | 340×340–430×430 | 4 | 3570–5020 | 60, 70 | 230 | 108 | 2 | STIR |
| DWI** | 2D-IR-DWI-EPI | 256×160–200×34–49 | 350×219–430×269 | 4 | 6290–9660 | 66, 70 | 220, 250 |  | 3/8/20 or 3/8/15*** | STIR |

FoV=field of view, TR=repetition time, TE=echo time, IR=inversion recovery time, DWI=diffusion-weighted imaging, GRE=gradient echo, SE=spin echo, EPI=echo-planar imaging, GBCA=gadolinium-based contrast agent, STIR=short T1 inversion recovery
\*Acquired before and during the five timepoints after intravenous GBCA injection (gadobutrol; Bayer, Leverkusen, Germany; 0.1 mmol/kg/body weight, injection speed=2 mL/s)
\*\*DWI scan was acquired using three b-values: 50, 750, and 1500 s/mm<sup>2</sup>
\*\*\*No. of averages for DWI indicates the number of averages for each b-value. The first set of values is for the acquisition performed on Skyra-Fit and the second set for that performed on MAGNETOM VIDA.

**Table 2:**
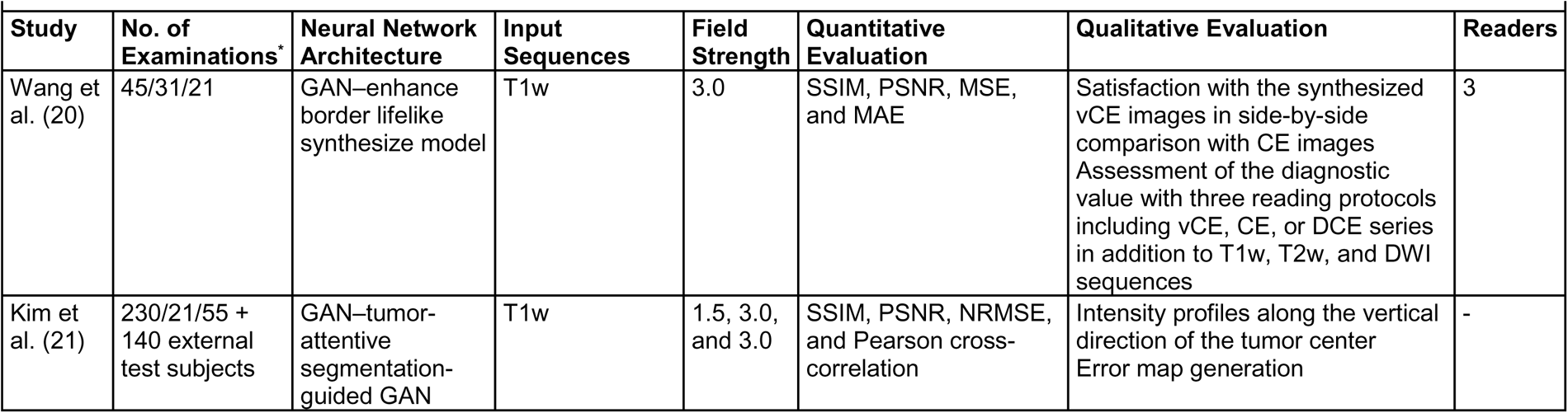
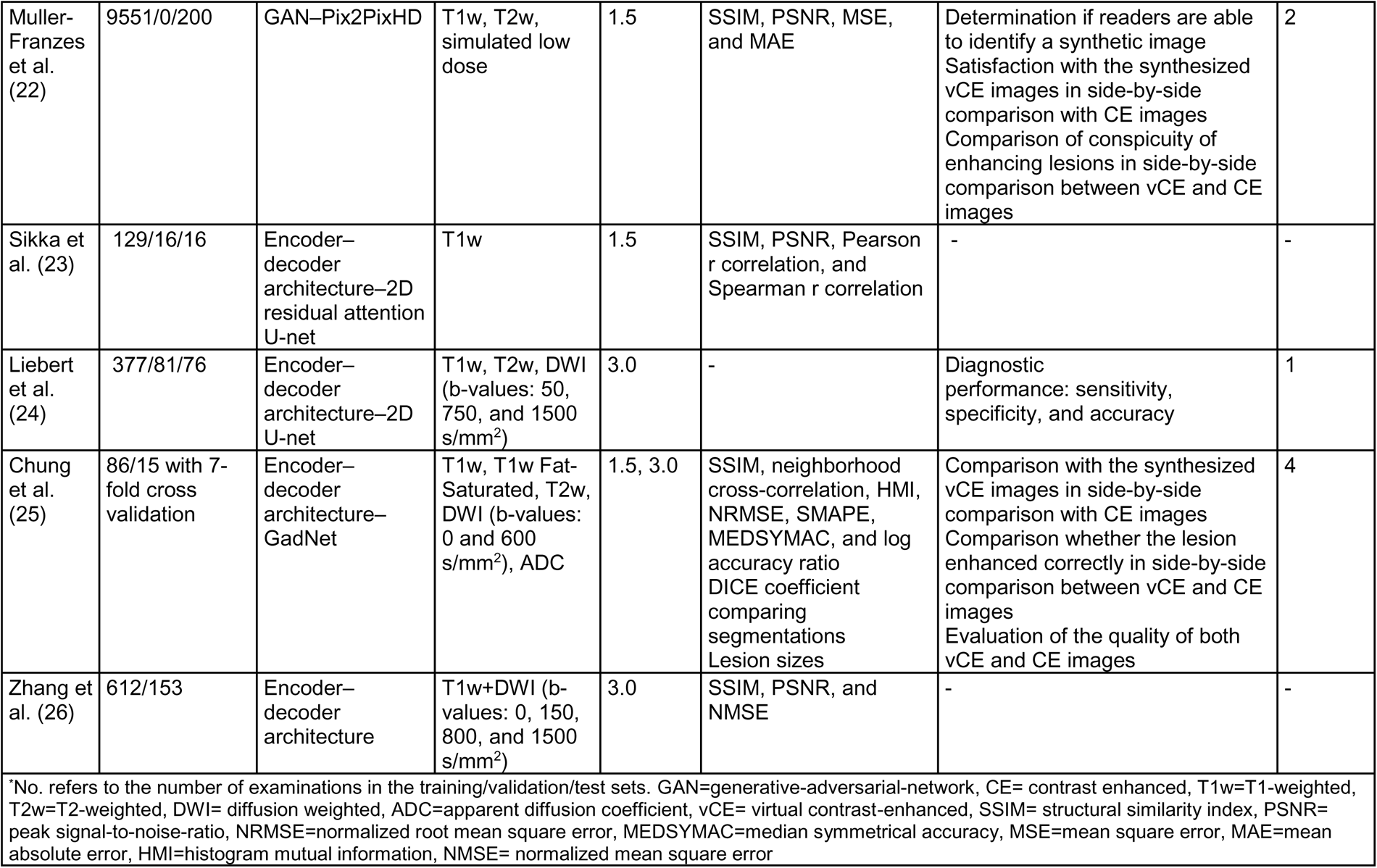
Literature Review.

| Table 2: Literature Review |  |  |  |  |  |  |  |
| --- | --- | --- | --- | --- | --- | --- | --- |
| Study | No. of Examinations* | Neural Network Architecture | Input Sequences | Field Strength | Quantitative Evaluation | Qualitative Evaluation | Readers |
| Wang et al. (20) | 45/31/21 | GAN–enhance border lifelike synthesizer model | T1w | 3.0 | SSIM, PSNR, MSE, and MAE | Satisfaction with the synthesized vCE images in side-by-side comparison with CE images<br>Assessment of the diagnostic value with three reading protocols including vCE, CE, or DCE series in addition to T1w, T2w, and DWI sequences | 3 |
| Kim et al. (21) | 230/21/55 + 140 external test subjects | GAN–tumor-attentive segmentation-guided GAN | T1w | 1.5, 3.0, and 3.0 | SSIM, PSNR, NRMSE, and Pearson cross-correlation | Intensity profiles along the vertical direction of the tumor center<br>Error map generation | - |
| Muller-Franzes et al. (22) | 9551/0/200 | GAN–Pix2PixHD | T1w, T2w, simulated low dose | 1.5 | SSIM, PSNR, MSE, and MAE | Determination if readers are able to identify a synthetic image<br>Satisfaction with the synthesized vCE images in side-by-side comparison with CE images<br>Comparison of conspicuity of enhancing lesions in side-by-side comparison between vCE and CE images | 2 |
| Sikka et al. (23) | 129/16/16 | Encoder–decoder architecture–2D residual attention U-net | T1w | 1.5 | SSIM, PSNR, Pearson r correlation, and Spearman r correlation | - | - |
| Liebert et al. (24) | 377/81/76 | Encoder–decoder architecture–2D U-net | T1w, T2w, DWI (b-values: 50, 750, and 1500 s/mm<sup>2</sup>) | 3.0 | - | Diagnostic performance: sensitivity, specificity, and accuracy | 1 |
| Chung et al. (25) | 86/15 with 7-fold cross validation | Encoder–decoder architecture–GadNet | T1w, T1w Fat-Saturated, T2w, DWI (b-values: 0 and 600 s/mm<sup>2</sup>), ADC | 1.5, 3.0 | SSIM, neighborhood cross-correlation, HMI, NRMSE, SMAPE, MEDSYMACE, and log accuracy ratio<br>DICE coefficient comparing segmentations<br>Lesion sizes | Comparison with the synthesized vCE images in side-by-side comparison with CE images<br>Comparison whether the lesion enhanced correctly in side-by-side comparison between vCE and CE images<br>Evaluation of the quality of both vCE and CE images | 4 |
| Zhang et al. (26) | 612/153 | Encoder–decoder architecture | T1w+DWI (b-values: 0, 150, 800, and 1500 s/mm<sup>2</sup>) | 3.0 | SSIM, PSNR, and NMSE | - | - |

\*No. refers to the number of examinations in the training/validation/test sets. GAN=generative-adversarial-network, CE= contrast enhanced, T1w=T1-weighted, T2w=T2-weighted, DWI= diffusion weighted, ADC=apparent diffusion coefficient, vCE= virtual contrast-enhanced, SSIM= structural similarity index, PSNR= peak signal-to-noise-ratio, NRMSE=normalized root mean square error, MEDSYMACE=median symmetrical accuracy, MSE=mean square error, MAE=mean absolute error, HMI=histogram mutual information, NMSE= normalized mean square error

## RESULTS

### Demographics and Clinical Findings

The demographics of the full data set and respective subsets are presented in Table 3. In the test cohort (n=187), n=67 (35.8%) cases had a malignant lesion (median size = 20.4 mm, 25^th^ – 75th percentile = 13.0 – 37.7 mm). Amongst the n = 67 malignant lesions the following histopathology was described: n = 11 (16%) DCIS, n = 7 (10%) ILCs, n = 1 mucinous carcinoma, n = 48 (71%) invasive breast cancer NST amongst n = 10/48 NST with associated DCIS.

Based on the clinical report n = 145 (77.5%) of the examinations had a target finding as defined in the methods (median size = 14.8mm, 25th–75th percentile = 7.1–26.4mm).

**Table 3:** Demographics of the Training/Validation and Independent Test Cohorts.

|  |  |  |  |  |
| --- | --- | --- | --- | --- |
|  | Entire dataset | Training | Validation | Independent Test Set |
| No. of patients | 869 (100%) | 606 (69.7%) | 76 (8.8%) | 187 (21.5%) |
| No. of examinations | 973 | 710 | 76 | 187 |
| Split ratio of examinations (%) | 100 | 73 | 7.8 | 19.2 |
| Age (year) | 52±12 | 50±12 | 52±12 | 51±14 |
| Routine BI-RADS score* |  |  |  |  |
| 0 | 23 (2.4%) | 20 (2.8%) | 0 (0%) | 3 (1.6%) |
| 1 | 3 (0.3%) | 2 (0.3%) | 0 (0%) | 1 (0.5%) |
| 2 | 461 (47.4%) | 330 (46.5%) | 42 (55.3%) | 89 (47.6%) |
| 3 | 73 (7.5%) | 49 (6.9 %) | 7 (9.2%) | 17 (9.1%) |
| 4 | 114 (11.7%) | 96 (13.5%) | 4 (5.3%) | 14 (7.5%) |
| 5 | 70 (7.2%) | 48 (6.8%) | 3 (4.0%) | 19 (10.2%) |
| 6 | 229 (23.5%) | 165 (23.2%) | 20 (26.3%) | 44 (23.5%) |
| *The BI-RADS score refers to the highest BI-RADS score given for an examination during routine clinical reading; age is presented as means and standard deviations. Percentages of BI-RADS scores are given as ratio of the respective dataset |  |  |  |  |

#### Experiment 1: Quantitative Analysis

Figure 2 shows the technical performance metrics of all 11 input combinations in the entire breast volume and segmented findings. Significant differences were found in all quantitative scores using the Friedmann test for both breast volume evaluation and target finding evaluations (all p<0.001). The means and standard deviations of the quantitative metrics and detailed post-hoc Nemanyi test results of multiple comparisons of the input sequence performances are presented in the Figure 3.

In the breast volume similarity metrics increased significantly when T2w sequences were added to the T1w sequences alone (SSIM = 87.06±2.57 vs 86.91±2.58, p=0.011, PSNR = 24.33±1.79 vs 24.18±1.82dB, p=0.011). However, while both error metrics decreased (NRMSE= 8.77±1.17 vs 8.91±1.20; MEDSYMAC = 2.02±0.92 vs 2.08±0.95 dB) only the NRMSE metric changed significantly (NRMSE p=0.011, MEDSYMAC p=1.0). The T1w sequences alone yielded significantly higher similarity metrics than the T2w (p=0.011) or both combination of the DWI (p=0.011) alone as well as both combination of just these two sequences (both combinations p=0.011). The full-breast volume similarity metrics did not significantly differ neither when the b50/b750 combination nor when the b50/b750/b1500 combination (SSIM = 87.00±2.55 p=1.0, PSNR = 22.82±1.70dB p=1.0) was added to the T1w sequence (SSIM = 87.00±2.55 for the combination of T1w, b50/b750/ b1500 p=1.0) when compared to the input of just T1w sequence. Significant improvement in regards to the similarity metrics could be observed when both T2w and different combinations of DWI sequences were added the T1w when compared just to the T1w input sequence (p=0.11 for both metrics and both comparisons).

When the metrics of the segmented target findings within the breast were considered, the SSIM significantly increased when multi-b-value DWI with ultra-high b-values of b1500 was added, from 50.25±22.84 (T1w) and 51.94±22.80 (T1w, T2w) to 63.08±20.16 (p=.011) and the MEDSYMAC decreasing from 13.92±5.19 (T1w) and 14.12±6.28 (T1w, T2w) to 11.01±5.07 (p=.011 for both). Adding T2w sequences to the combination of T1w, b50/b750/b1500 did not significantly influence the quantitative metrics (p=1.0) in any evaluation. Further numerical results of all sequence combinations are shown in Supplement Material Table 1.

**Figure 2:**
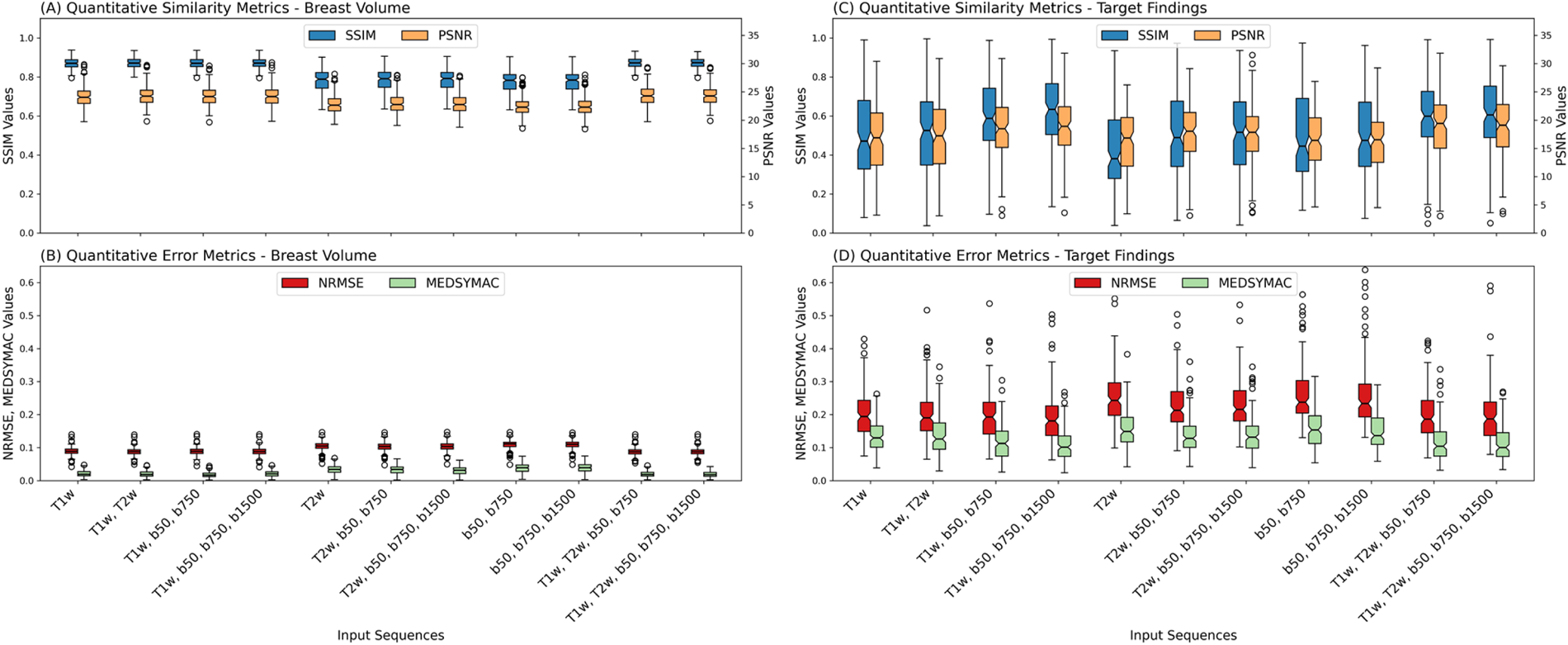
Box charts for the quantitative similarity (SSIM and PSNR) and error (NRMSE and MEDSYMAC) metrics for the full cohort (a and b, respectively) and only the enhancing lesions (c and d, respectively). A significant difference in the SSIM and PSNR could be observed in the breast volume between the input combinations with and without a T1w image acquisition. The similarity and error metrics were significantly higher and significantly lower in the target findings, respectively, when compared with the input sequence combinations including both T1w image and DWI acquisitions and combinations that missed either of those acquisitions. T1w=T1-weighted, T2w=T2-weighted, b50=DWI acquisition with a b-value of 50 s/mm^2^, b750=DWI acquisition with a b-value of 750 s/mm^2^, b1500=DWI acquisition with a b-value of 1500 s/mm^2^, SSIM= structural similarity index, PSNR= peak signal-to-noise-ratio, NRMSE=normalized root mean square error, MEDSYMAC=median symmetrical accuracy

**Figure 3:**
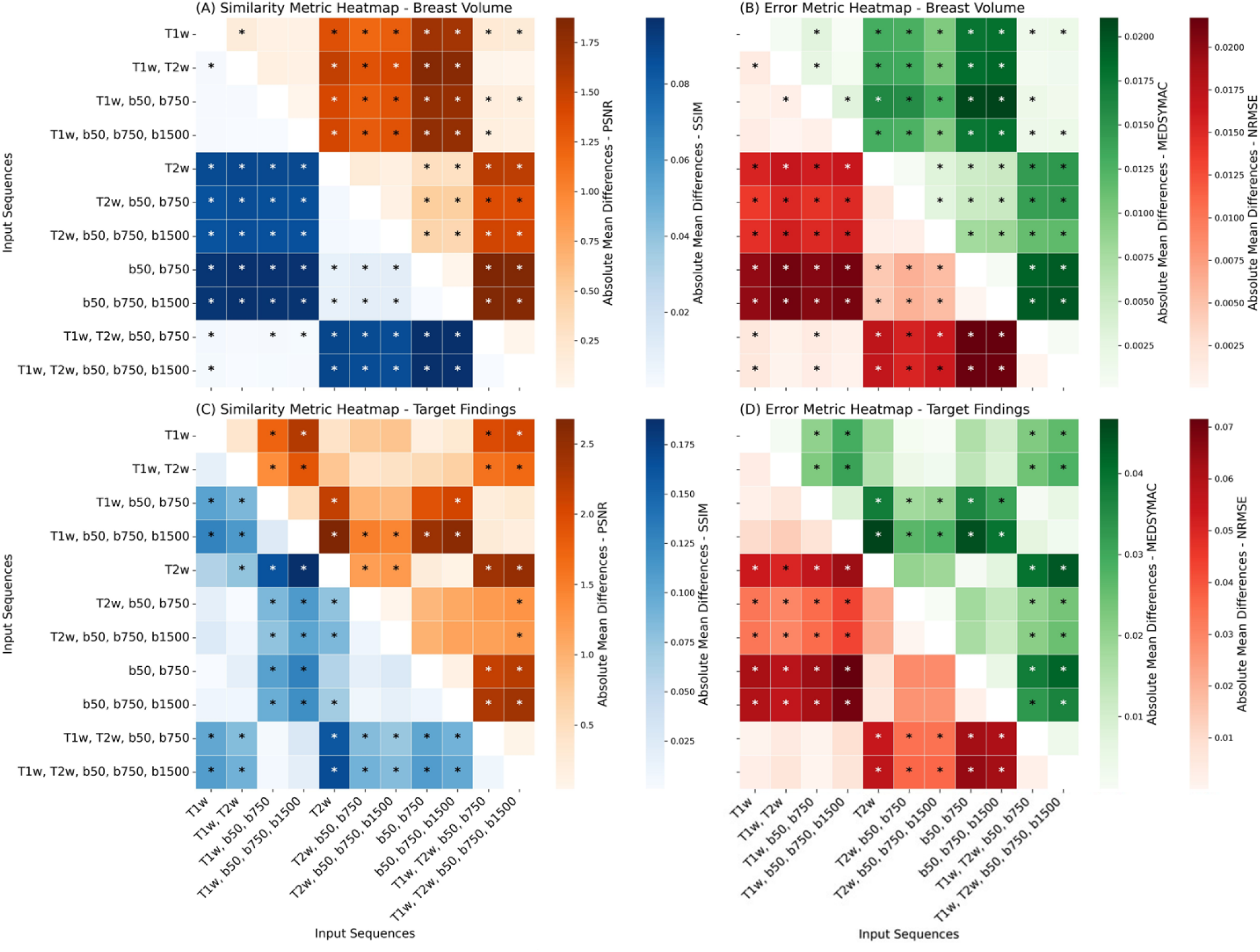
Correlation plot showing the mean absolute differences in the median reading scores for the quantitative similarity (a and c) and error (b and d) metrics in the entire breast volume (a and b) and target findings (c and d). T1w=T1-weighted, T2w=T2-weighted, b50=DWI acquisition with a b-value of 50 s/mm^2^, b750=DWI acquisition with a b-value of 750 s/mm^2^, b1500=DWI acquisition with a b-value of 1500 s/mm^2^, Post-Contrast=subtraction of the second post-contrast phase of a DCE image acquisition. *p<.05, **p<.01, ***p<.001 Target findings refer to the findings within the examination that could be both benign or malignant and non-mass or mass enhancement as well as findings not enhancing (e.g., cysts) but morphologically delineated from healthy fibro-glandular tissue.

#### Experiment 2: Qualitative Analysis

Four cases are demonstrated in Figure 4A–D. N=93/187 (49.7%) of the independent test cases were identified as showing an enhancing lesion (n=61/93, 65.6%) or NME (n=32/93, 34.4%) by at least two readers (further referred to as enhancing lesions/NMEs). The box plots for the qualitative reading are presented in Figure 5 for the entire patient cohort and the cases with enhancing lesions/NMEs. Significant differences were found in all qualitative scores using the Friedmann test for both the full cohort and cases with enhancing lesions/NMEs (all p-values: <0.001, highest p-value = 6.18e-68 for contrast/SNR satisfaction in the enhancing lesions). The post-hoc Nemanyi test results are presented in Figure 6. The box plots for the qualitative scores among the readers are presented in the Supplement Material Figure 2 and 3.

The input sequence combinations not containing a T1w sequence showed significantly lower median values for the overall image quality and image sharpness for both the full cohort and cases with enhancing lesions/NMEs (highest p-vale: 0.012, Figure 6). The best performing input sequence combination in regards to both image sharpness and the diagnostic image quality was the combination of T1w, T2w, b50/b750. For the full cohort all combination which included any DWI acquisition (either b50/b750 or b50/b750/b1500) showed no significant difference (all p-values=1.00) in regards or the lesion/NME conspicuity when compared to original Post-Contrast images. The best performing input combination in this regard was the combination of b50/b750/b1500.

In regards to contrast satisfaction only combination which included both DWI acquisitions and T1w acquisitions showed no significant difference (p=1.0) when compared to the original Post-Contrast images for the full cohort. However, in the subgroup analysis of the enhancing lesions/NME’s it could be noted that a significant difference from the Post-Contrast could be observed for all input combinations (highest p=0.012). The best performing input sequence combination in regards to the contrast satisfaction was the combination of T1w, T2w b50/b750/b1500 for both the full cohort and for the enhancing lesions/NME’s.

The combinations best-performing in regards to quantitative values (T1w, b50/b750/b1500) didn’t significantly differed from the original Post-Contrast images in regards to all of the reading features (p=1.0).

Both lesion/NME conspicuity and contrast satisfaction improved when T1w image acquisition was used together with the ultra-high b-value (b50/b750/b1500) compared with those when only a low b-value (b50/b750) was utilized, although the difference was not significant (p=1.0 for both with and without inclusion of T2w). Example cases showing such visual improvement of the lesion conspicuity by the inclusion of the b1500 are shown in Figures 4C and 4D.

Table 4 shows the three best performing input combinations in regards to each of the reading features. Based on this evaluation it can be noted that the combination of T1w, T2w, b50/b750/1500 was the only combination which appears among the top three combinations for all of the four reading features whilst the combination of T1w, b50/b750/1500 appeared three times on the list, however providing a higher lesion enhancement score than the combination including an additive T2w sequence.

An individual in depth exploration of these combinations a) T1w, b50/b750/1500 and b) T1w, T2w, b50/b750/1500 was therefore performed as the following:

For both input combinations, the analysis revealed that n=93/93 of the enhancing lesions were identified to provide at least a minimal enhancement score (score 1) by at least two of the three readers on the vCE data. However, for input combination T1w, b50/b750/1500, n=15/93 (n=6 malignant, n=9 benign) (16.1%) of these lesions reached less than 50% of the maximal enhancement score given on CE images and n=6/93 (6.5%) lesions were attributed a minimal enhancement score of 1, which included n=1 malignant (breast cancer NST, cT1) and n=5 benign lesions (scar tissue (n=1), mastopathy (n=2), fibroadenoma (n=2). Amongst these n=6 lesions n=5 cases were masses and one case appeared as NME. Lesion size evaluation revealed n=3 of those cases to be under <10 mm of size, n=1 with a size between 10 and 20 mm and n=2 with a size above 20 mm. In contrast, the combination of T1w, T2w, b50/b750/1500, demonstrated n=8/93 lesions (8.6%) with an enhancement score of 1, which included the beforementioned malignant case and two additive malignant cases (both breast cancer cases with an NST) and the beforementioned benign cases. For this input combination among the n=8 lesions showing a minimal enhancement score of 1, n=7 cases were masses and one case appeared as NME. Lesion size evaluation revealed n=3 of those cases to be under <10 mm of size, n=2 with <20mm of size and n=3 with a size above 20 mm.

**Table 4:** Best performing combinations.

| **Table 4: Best performing combinations** |  |  |  |
| --- | --- | --- | --- |
| Reading Feature | 1 | 2 | 3 |
| Diagnostic Image Quality | T1w, T2w, b50, b750 | **T1w, T2w, b50, b750, b1500** | T1w, b50, b750, b1500 |
| Image Sharpness | T1w, T2w, b50, b750 | **T1w, T2w, b50, b750, b1500** | T1w |
| Satisfaction with SNR/Contrast | **T1w, T2w, b50, b750, b1500** | T1w, b50, b750, b1500 | T1w, T2w, b50, b750 |
| Lesion Enhancement | b50, b750, b1500 | T1w, b50, b750, b1500 | **T1w, T2w, b50, b750, b1500** |
| The only input combination appearing for all four reading features among is marked with bold.<br>T1w=T1-weighted, T2w=T2-weighted, b50 – DWI with a b-value of 50 s/mm<sup>2</sup>, b750 – DWI with a b-value of 750 s/mm<sup>2</sup>, b1500 – DWI with a b-value of 1500 s/mm<sup>2</sup>, SNR – signal-to-noise ratio |  |  |  |

**Figure 4A:**
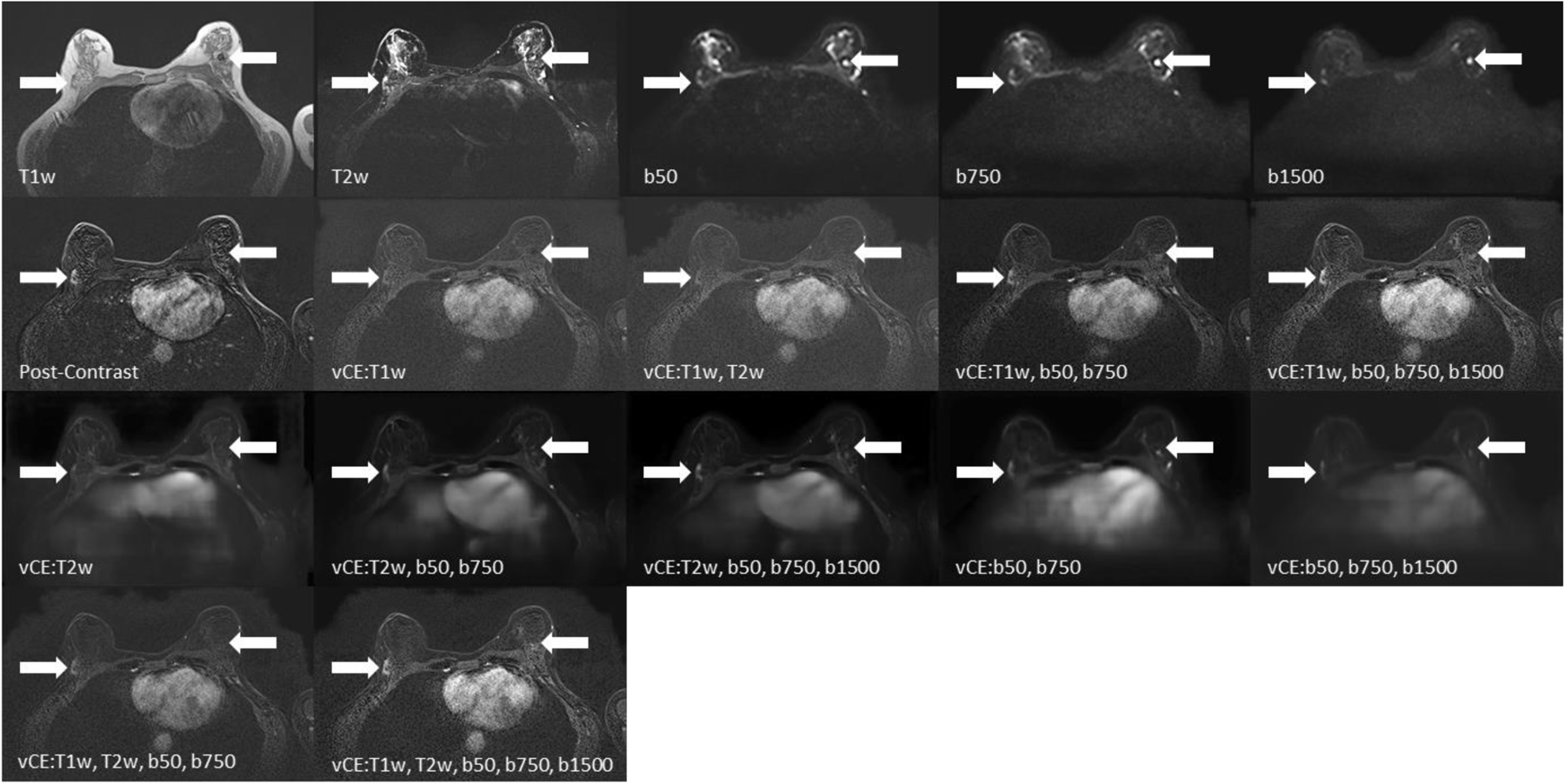
Diagnostic breast MRI slices that were read by the three readers showing the input sequences, post-contrast-enhanced image acquisition, and 11 vCE versions. Patient in their 60s, with a lesion in the right breast (16.2 mm, white arrow) enhancing in the GBCA-enhanced post-contrast subtraction image (“Post-Contrast”). The lesion does not enhance in the vCE images that did not include any DWI sequences as input. Histopathology confirmed a breast carcinoma (NST) with DCIS (ductal carcinoma in situ). An additive finding of a complex cyst is demonstrated in this case in the left breast with a high signal in the b=1500s/mm^2^ b-value DWI and a heterogeneous signal in the T1w sequence. In the vCE data, the carcinoma shows contrast-enhancement whilst the complex cyst does not simulate a contrast-agent uptake. T1w=T1-weighted, T2w=T2-weighted, b50, b750, b1500 = DWI acquisitions with b-values= 50, 750 and 1500 s/mm^2^, Post-Contrast=subtraction of the 2nd post-contrast phase of a dynamic contrast enhanced image acquisition, vCE=virtual contrast-enhanced image generated using the respective combination of input sequences.

**Figure 4B:**
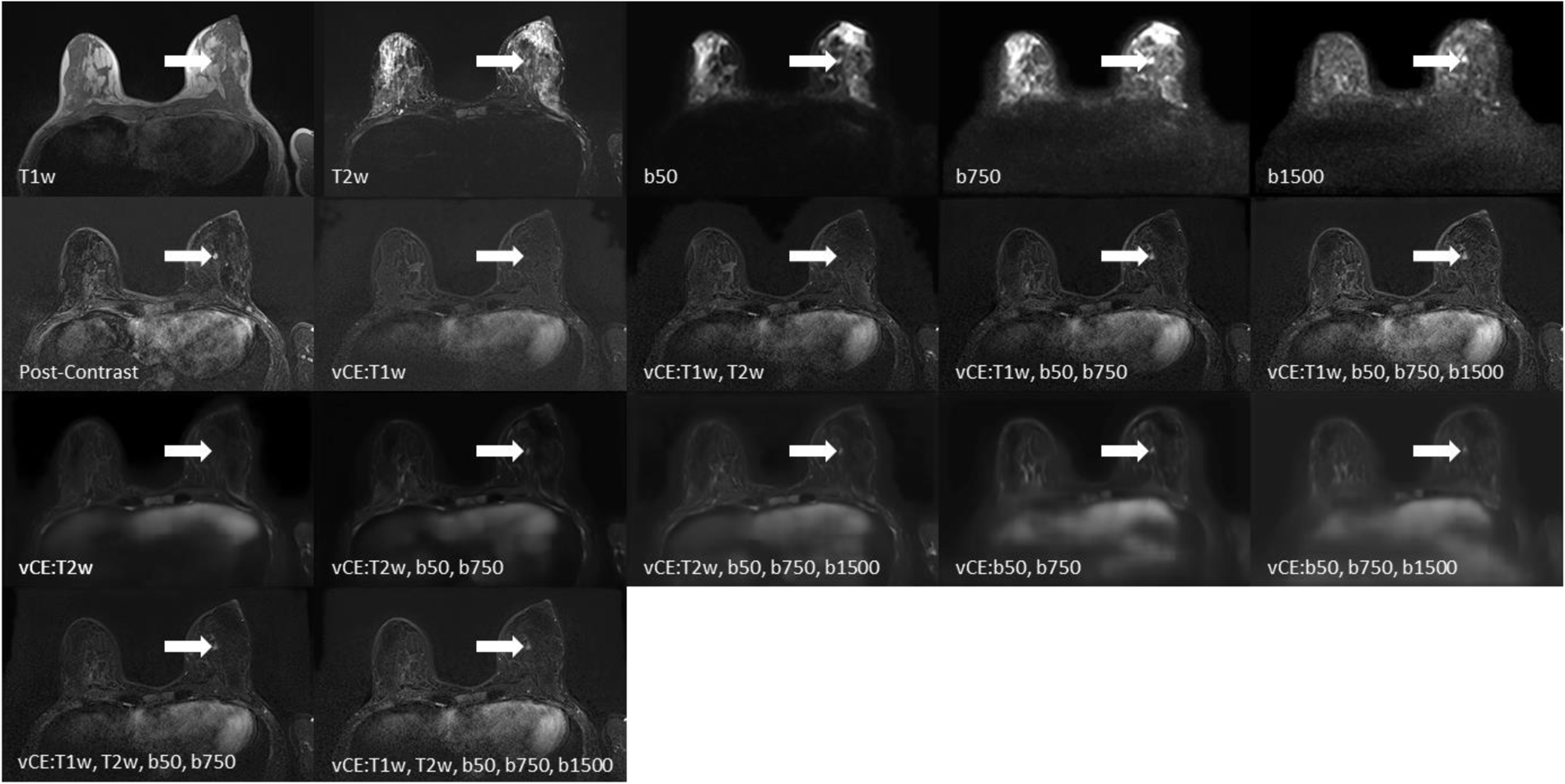
Diagnostic breast MRI slices that were read by the three readers showing the input sequences, post-contrast-enhanced image acquisition, and 11 virtual contrast-enhanced versions. Scans of a patient in their 40’s show a lesion (5.7-mm, white arrow) enhancing in the GBCA-enhanced post-contrast subtraction image (“Post-Contrast”). The lesion does not enhance in the vCE images that did not include any DWI sequences as input. Histopathology confirmed DCIS (ductal carcinoma in situ). T1w=T1-weighted, T2w=T2-weighted, b50, b750, b1500 = DWI acquisitions with b-values= 50, 750 and 1500 s/mm^2^, Post-Contrast=subtraction of the 2nd post-contrast phase of a dynamic contrast enhanced image acquisition, vCE=virtual contrast-enhanced image generated using the respective combination of input sequences.

**Figure 4C:**
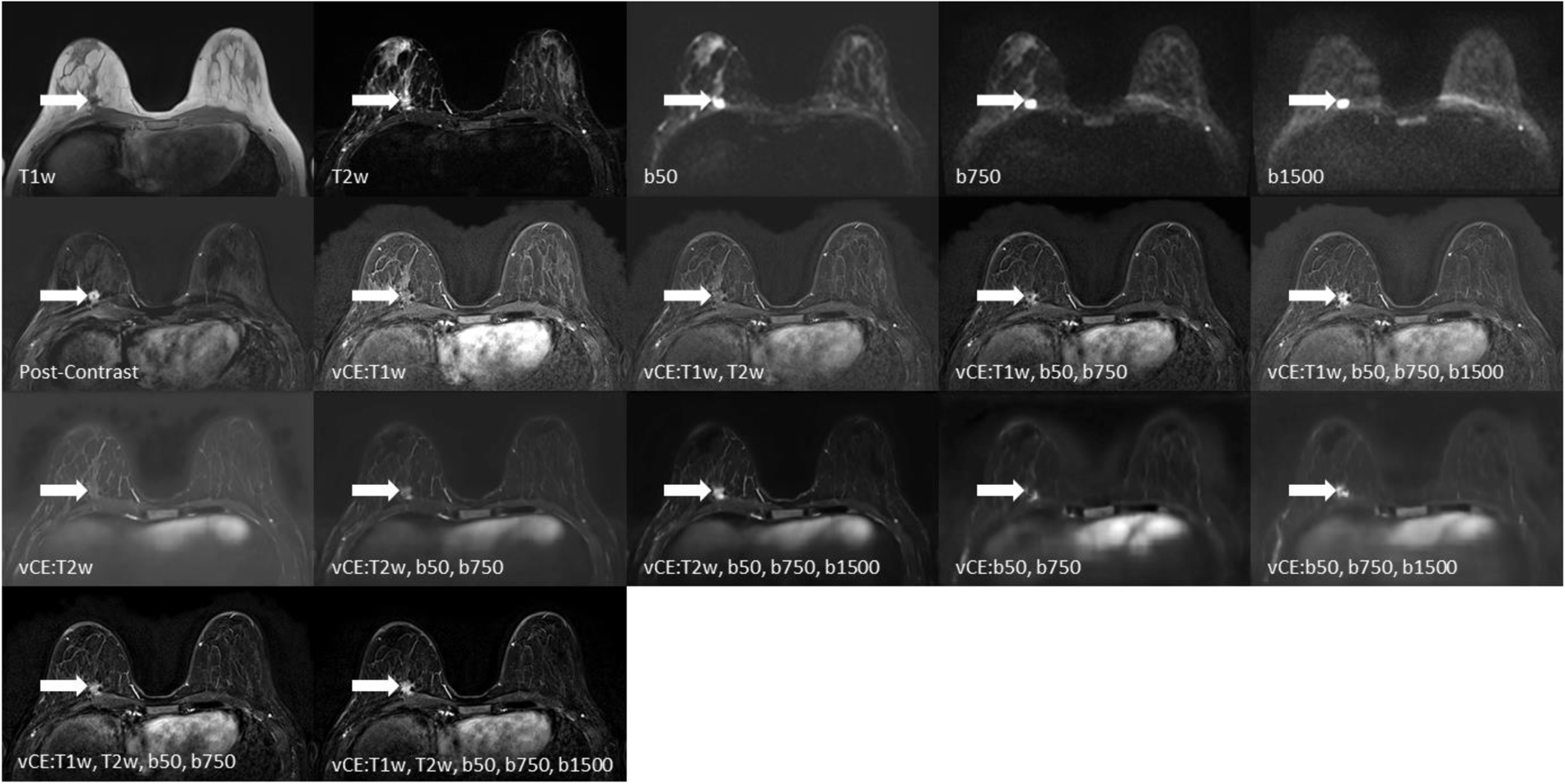
Diagnostic breast MRI slices that were read by the three readers showing the input sequences, post-contrast-enhanced image acquisition, and 11 virtual contrast-enhanced versions. The scans show a patient in their 40’s (13 mm, white arrow) showing a lesion in the right breast enhancing in the GBCA-enhanced post-contrast subtraction image (“Post-Contrast”). The lesion barely enhances in the vCE images that did not include any DWI sequence as input. A more pronounced enhancement could be observed in the vCE versions including a b1500 input sequence as compared with the vCE approaches including only b50 and b750. Histopathology confirmed breast carcinoma (NST). T1w=T1-weighted, T2w=T2-weighted, b50, b750, b1500 = DWI acquisitions with b-values= 50, 750 and 1500 s/mm^2^, Post-Contrast=subtraction of the 2nd post-contrast phase of a dynamic contrast enhanced image acquisition, vCE=virtual contrast-enhanced image generated using respective combination of input sequences.

**Figure 4D:**
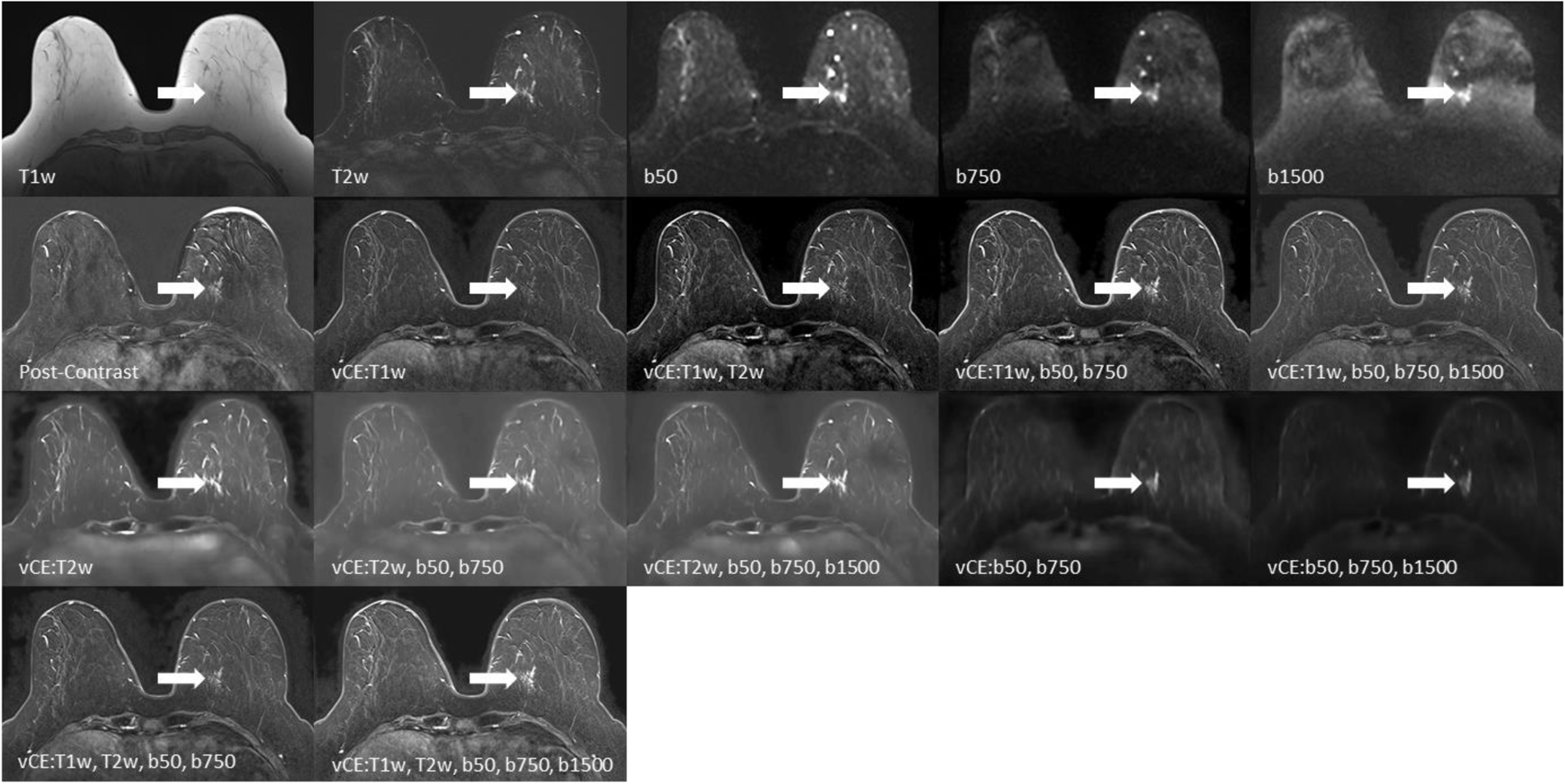
Diagnostic breast MRI slices that were read by the three readers showing the input sequences, post-contrast-enhanced image acquisition, and 11 virtual contrast-enhanced versions. The scans show patient in their 60’s showing a non-mass enhancement (NME) in the right breast (21.4 mm area, white arrow) enhancing lightly in the GBCA-enhanced post-contrast subtraction image. The NME does not enhance distinctly in the vCE images that did not include any DWI sequence as input. A more pronounced enhancement could be observed in the vCE versions including a b1500 input sequence as compared with the vCE training only including b50, b750 DWI sequences. Histopathology confirmed invasive breast cancer. T1w=T1-weighted, T2w=T2-weighted, b50, b750, b1500 = DWI acquisitions with b-values= 50, 750 and 1500 s/mm^2^, Post-Contrast=subtraction of the 2nd post-contrast phase of a dynamic contrast enhanced image acquisition, vCE=virtual contrast-enhanced image generated using the respective combination of input sequences.

**Figure 5:**
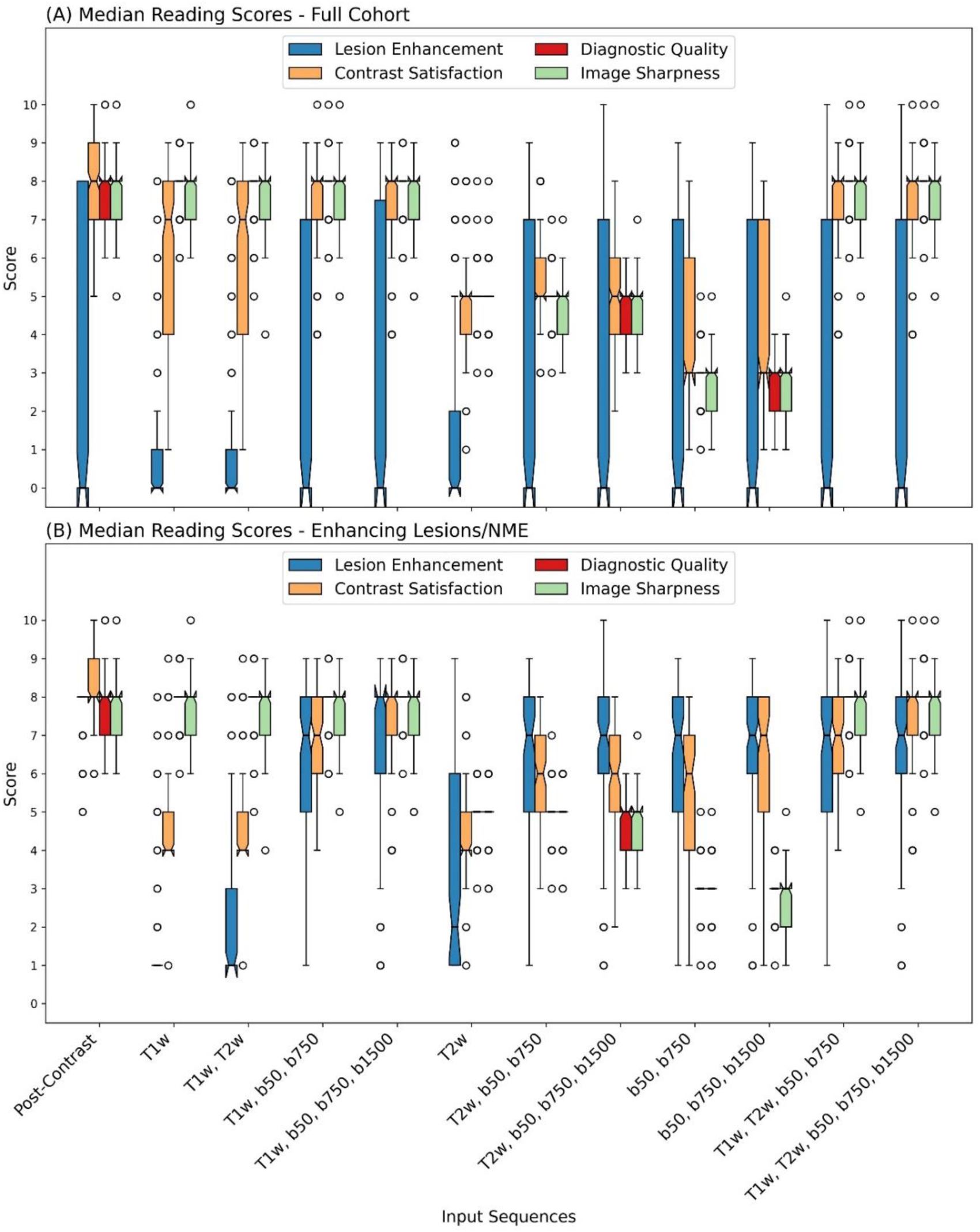
Box plots for the median reading scores among the three readers for the full cohort and the cases with enhancing lesions/NMEs in the GBCA-enhanced post-contrast subtraction image. Significantly lower lesion/NME enhancement could be observed when no DWI acquisitions were included in the input sequence combination. A lower contrast satisfaction score could be noted in the enhancing lesions/NMEs with no b1500 acquisition than with inclusion (e.g., T1w, b50, b750 vs. T1w, b50, b705, b1500). All input combinations that did not include a T1w sequence showed a lower score in the diagnostic quality and image sharpness. T1w=T1-weighted, T2w=T2-weighted, b50=DWI acquisition with a b-value of 50 s/mm^2^, b750=DWI acquisition with a b-value of 750 s/mm^2^, b1500=DWI acquisition with a b-value of 1500 s/mm^2^, Post-Contrast=subtraction of the second post-contrast phase of a DCE image acquisition.

**Figure 6:**
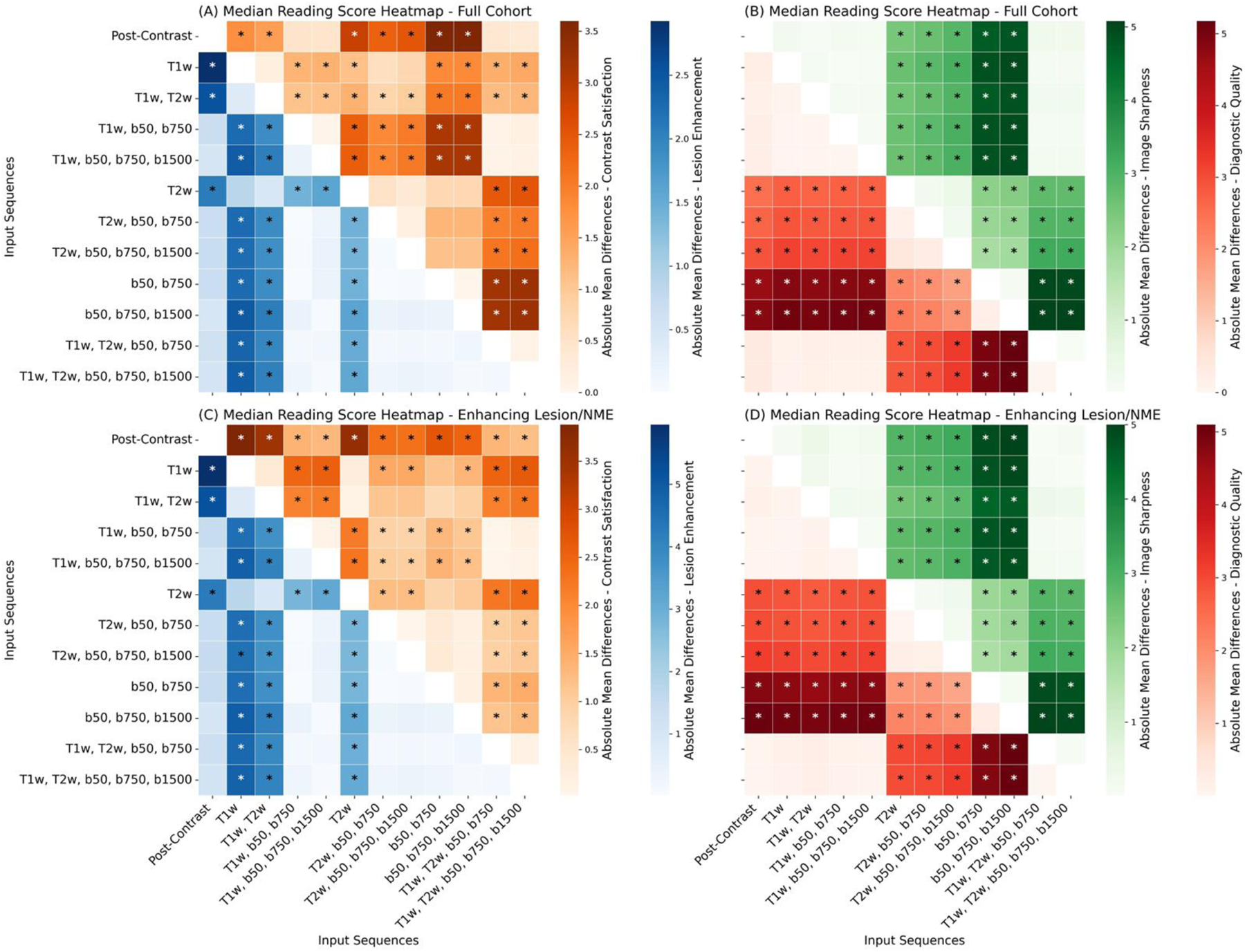
Correlation plot showing the mean absolute differences in the median reading scores for the full cohort (a and b) and the cases with enhancing lesions/NMEs in the post-contrast subtraction image after GBCA injection (c and d). Significance in the difference of the median reading score between the respective methods are marked with stars. T1w=T1-weighted, T2w=T2-weighted, b50=DWI acquisition with a b-value of 50 s/mm^2^, b750=DWI acquisition with a b-value of 750 s/mm^2^, b1500=DWI acquisition with a b-value of 1500 s/mm^2^, Post-Contrast=subtraction of the second post-contrast phase of a DCE image acquisition. *p<.05, **p<.01, ***p<.001

## DISCUSSION

This study demonstrated that the performance of virtual contrast-enhanced breast MRI neural networks significantly depended on the acquisition sequences provided as input during training. The highest overall performance regarding the conspicuity of lesions/NMEs depicted in gadolinium-based contrast agent-enhanced image acquisition both qualitatively and quantitatively was observed when multi-b-value diffusion-weighted imaging was combined with an ultra-high b-value with high-resolution morphologic T1-weighted image acquisition.

Our data further demonstrate the relevance of visual readings and clinical target-focused metrics, since the metrics based on the full image volume were much less influenced by the input variations than the metrics specifically considering the target findings within the breast tissue (both enhancing and non-enhancing findings).

Using non-enhanced MRI acquisitions for deriving vCE images is an emerging field of research in breast MRI (20–25). Different approaches have been suggested, with many studies focusing on morphologic acquisitions during neural network training. T1w image acquisitions partially combined with T2w image acquisitions depicting morphologic anatomy thus have built the foundation for the studies by Wang et al., Kim et al., Sikka et al., and Mueller-Franzes et al. (20–23,26). Mueller-Franzes et al. (22) conducted the largest study on this vCE approach. Their results suggest a limited capability of vCE imaging using morphologic sequences alone in depicting enhancing lesions, which agrees with our data.

Our data also suggest a significant contribution of DWI acquisitions to the ability of vCE imaging to correctly depict enhancing lesions/NMEs. This finding is supported by the reports of Chung et al. (25) and Zhang et al.(26) However until now, it remained unclear, if the DWI acquisition schemes, e.g. similar to the one recommended by the EUSOBI (34) or including an additive ultra-high b-value (35) can influence neural networks trained to derive vCE imaging data. Our results suggest that the presence of an ultra-high b-value might improve lesion/NME conspicuity/enhancement on edge cases; however, the differences did not reach significance for our sample size. This can be caused by the fact that, if acquired with a sufficient image quality, ultra-high b-value DWI scans can help detect suspicious lesions in the breast (35,36) and within healthy fibro-glandular tissue, which commonly loses signal beyond a b-value of approximately 1250 s/mm^2^. The vCE approach in our study focused on reflecting the post-contrast acquisition performed about 110–130 s after GBCA injection. Accordingly, the contrast enhancement patterns at this timepoint were influenced not only by perfusion fractions but also by more complex tissue alterations associated with lesion growth such as microstructural heterogeneity and tissue complexity, which may be partially reflected in ultra-high b-value image acquisitions (37,38).

The quantitative similarity (SSIM and PSNR) and error metrics (NRMSE and MEDSYMAC) of the full images demonstrated only minor differences compared with the results of the reader studies and focused analyses of the target findings. Notably, even the images trained on T1w images alone provided overall high similarity and low error values in the entire breast volume. However, false negatives regarding lesions/NMEs enhancement were consistently found with this approach by all three readers. Such situation can be attributed to the fact that suspicious lesions/NMEs constitute a substantially small fraction of images; hence, during averaging of the similarity and error metrics over the whole breast volume, the missing enhancement does not influence the final quantitative metrics.

Patient characteristics might additively influence the results and limit the generalizability of vCE image generation models. Chung et al. (25) exclusively included patients with biopsy-proven invasive breast cancer with a mean lesion size of 24 mm. This cohort matches that in the study by Zhang et al.(26), but who did not disclose the lesion size in detail. In contrast, Mueller-Franzes et al. (22) investigated women undergoing breast MRI for screening with a consecutively smaller median lesion size of about 15 mm. With a median size of all findings of 14.5 mm and a median malignant lesion size of 20.4 mm, our study is in between these two studies. Our study further included a diverse clinical real-life spectrum of women undergoing breast MRI in a tertiary hospital. We did not further select patient subgroups to avoid selection bias, influencing the network and allowing for an inclusion of a natural distribution of lesion sizes, lesion characteristics, and clinical indications for breast MRI.

As this field of research is yet in its infancy, many questions and challenges must be addressed, including the limitations of our study. Although our study included different MRI scanner types and a real-world spectrum of clinical cases encountered in breast MRI in the training and test sets, further research is required to understand limits regarding generalizability, robustness, and clinical applicability. This expands to sequence setting variations, vendors, and field strengths. Moreover, we investigated only a single encoder-decoder neural network architecture but future studies should evaluate other neural network architecture types such as other encoder decoder architectures, GAN’s or transformers. Next, diagnostic accuracy was not evaluated; “soft” criteria of lesion/NME conspicuity and image quality were rather selected, as previously described by Mueller-Franzes et al. and Chung et al., together with quantitative metrics. Thus, no conclusion could be drawn based on our data about the potential clinical applicability of the approach or about potentially relevant influencing clinical factors, especially with regards to the influence of lesion size on lesion visualization. Our study indicates, that some lesions show a significantly lower virtual enhancement as compared to GBCA-enhanced subtractions. This was the case even in input combination considered preferable for this task (input sequences: T1w, b50/b750/1500) with about 16.1% of the lesions demonstrating an enhancement score <50% of the GBCA-enhanced subtractions and n=6/92 lesions demonstrating only a minimal enhancement score of 1 – amongst one malignant lesion. Analyzing those cases however did not reveal a clear association to characteristics of histopathology, enhancement pattern (NME) or lesion size – yet a slightly higher scoring variance was observed for smaller lesions. It needs to be considered that our study sample size was not powered to investigate this dependency with a sufficient validity. Interestingly, while slightly improving overall image quality, adding the T2w sequence to the input tripled low-enhancing scores in malignant cases potentially suggesting fluid-overweighting in the input to impact the neuronal network leading to misinterpretation of enhancing lesions towards cystic findings. Larger-scale studies with multicentric designs and blinded readings are therefore necessary to determine the potential clinical application (e.g., in intermediate-risk screening). Further, although a broad spectrum of input sequences including their sub-settings (different b-value combinations) was explored, no conclusion could be drawn about potentially relevant complementary contrasts such as chemical exchange saturation transfer (39) or magnetic resonance fingerprinting.

In conclusion, a neural network for generating virtual contrast-enhanced images performed best when fed with a multiparametric unenhanced breast MRI protocol, which included both high-resolution morphologic information and multi-b-value diffusion-weighted imaging with ultra-high b-value image acquisition. Further research is needed to more comprehensively explore virtual contrast-enhanced approaches before assessing their clinical applicability.

## Data Availability

Original image data used in this work are not publicly available to preserve individuals privacy under the European General Data Protection Regulation. The institution handling this data is the Institute of Radiology University Hospital Erlangen.

## Abbreviations

CE: contrast enhanced
vCE: virtual contrast enhanced
SSIM: structural similarity index
PSNR: peak signal-to-noise-ratio
NRMSE: normalized root mean square error
MEDSYMAC: median symmetrical accuracy
NME: non-mass-enhancement
b50: diffusion weighted imaging acquisition with a b-value of 50 s/mm^2^
b750: diffusion weighted imaging acquisition with a b-value of 750 s/mm^2^
b1500: diffusion weighted imaging acquisition with a b-value of 1500 s/mm^2^

## REFERENCES

1. Mann RM, Cho N, Moy L. Breast MRI: State of the Art. Radiology 2019;292(3):520–536. doi: 10.1148/radiol.2019182947

2. Mann RM, Kuhl CK, Moy L. Contrast-enhanced MRI for breast cancer screening. Journal of Magnetic Resonance Imaging 2019;50(2):377–390. doi: 10.1002/jmri.26654

3. Mann RM, Athanasiou A, Baltzer PAT, Camps-Herrero J, Clauser P, Fallenberg EM, Forrai G, Fuchsjager MH, Helbich TH, Killburn-Toppin F, Lesaru M, Panizza P, Pediconi F, Pijnappel RM, Pinker K, Sardanelli F, Sella T, Thomassin-Naggara I, Zackrisson S, Gilbert FJ, Kuhl CK, European Society of Breast I. Breast cancer screening in women with extremely dense breasts recommendations of the European Society of Breast Imaging (EUSOBI). Eur Radiol 2022;32(6):4036–4045. doi: 10.1007/s00330-022-08617-6

4. Bakker MF, de Lange SV, Pijnappel RM, Mann RM, Peeters PHM, Monninkhof EM, Emaus MJ, Loo CE, Bisschops RHC, Lobbes MBI, de Jong MDF, Duvivier KM, Veltman J, Karssemeijer N, de Koning HJ, van Diest PJ, Mali WPTM, van den Bosch MAAJ, Veldhuis WB, van Gils CH. Supplemental MRI Screening for Women with Extremely Dense Breast Tissue. New England Journal of Medicine 2019;381(22):2091–2102. doi: 10.1056/NEJMoa1903986

5. Kuhl CK, Schmutzler RK, Leutner CC, Kempe A, Wardelmann E, Hocke A, Maringa M, Pfeifer U, Krebs D, Schild HH. Breast MR Imaging Screening in 192 Women Proved or Suspected to Be Carriers of a Breast Cancer Susceptibility Gene: Preliminary Results. Radiology 2000;215(1):267–279. doi: 10.1148/radiology.215.1.r00ap01267

6. Griebsch I, Brown J, Boggis C, Dixon A, Dixon M, Easton D, Eeles R, Evans D, Gilbert FJ, Hawnaur J. Cost-effectiveness of screening with contrast enhanced magnetic resonance imaging vs X-ray mammography of women at a high familial risk of breast cancer. British journal of cancer 2006;95(7):801–810.

7. Taneja C, Edelsberg J, Weycker D, Guo A, Oster G, Weinreb J. Cost effectiveness of breast cancer screening with contrast-enhanced MRI in high-risk women. Journal of the American College of Radiology 2009;6(3):171–179.

8. Plaza M, Cole D, Sanchez-Gonzalez MA, Starr CJ. Patient Throughput Times for Supplemental Breast Cancer Screening Exams. Archives of Breast Cancer 2021:21–28.

9. Borthakur A, Weinstein SP, Schnall MD, Conant EF. Comparison of Study Activity Times for “Full” versus “Fast MRI” for Breast Cancer Screening. J Am Coll Radiol 2019;16(8):1046–1051. doi: 10.1016/j.jacr.2019.01.004

10. Tollens F, Baltzer PA, Dietzel M, Schnitzer ML, Schwarze V, Kunz WG, Rink J, Rübenthaler J, Froelich MF, Schönberg SO. Economic potential of abbreviated breast MRI for screening women with dense breast tissue for breast cancer. European radiology 2022;32(11):7409–7419.

11. Tollens F, Baltzer PA, Dietzel M, Rübenthaler J, Froelich MF, Kaiser CG. Cost-effectiveness of digital breast tomosynthesis vs. abbreviated breast MRI for screening women with intermediate risk of breast cancer—how low-cost must MRI be? Cancers 2021;13(6):1241.

12. Mann RM, van Zelst JC, Vreemann S, Mus RD. Is ultrafast or abbreviated breast MRI ready for prime time? Current Breast Cancer Reports 2019;11:9–16.

13. McDonald RJ, Weinreb JC, Davenport MS. Symptoms Associated with Gadolinium Exposure (SAGE): A Suggested Term. Radiology 2022;302(2):270–273. doi: 10.1148/radiol.2021211349

14. Kleesiek J, Morshuis JN, Isensee F, Deike-Hofmann K, Paech D, Kickingereder P, Köthe U, Rother C, Forsting M, Wick W, Bendszus M, Schlemmer HP, Radbruch A. Can Virtual Contrast Enhancement in Brain MRI Replace Gadolinium?: A Feasibility Study. Invest Radiol 2019;54(10):653–660. doi: 10.1097/rli.0000000000000583

15. Chen C, Raymond C, Speier W, Jin X, Cloughesy TF, Enzmann D, Ellingson BM, Arnold CW. Synthesizing MR image contrast enhancement using 3D high-resolution ConvNets. IEEE Transactions on Biomedical Engineering 2022.

16. Haase R, Pinetz T, Bendella Z, Kobler E, Paech D, Block W, Effland A, Radbruch A, Deike-Hofmann K. Reduction of Gadolinium-Based Contrast Agents in MRI Using Convolutional Neural Networks and Different Input Protocols: Limited Interchangeability of Synthesized Sequences With Original Full-Dose Images Despite Excellent Quantitative Performance. Invest Radiol 2023;58(6):420–430. doi: 10.1097/RLI.0000000000000955

17. Calabrese E, Rudie JD, Rauschecker AM, Villanueva-Meyer JE, Cha S. Feasibility of simulated postcontrast MRI of glioblastomas and lower-grade gliomas by using three-dimensional fully convolutional neural networks. Radiology: Artificial Intelligence 2021;3(5):e200276.

18. Mallio CA, Radbruch A, Deike-Hofmann K, van der Molen AJ, Dekkers IA, Zaharchuk G, Parizel PM, Beomonte Zobel B, Quattrocchi CC. Artificial Intelligence to Reduce or Eliminate the Need for Gadolinium-Based Contrast Agents in Brain and Cardiac MRI: A Literature Review. Invest Radiol 2023. doi: 10.1097/RLI.0000000000000983

19. Gong E, Pauly JM, Wintermark M, Zaharchuk G. Deep learning enables reduced gadolinium dose for contrast-enhanced brain MRI. J Magn Reson Imaging 2018;48(2):330–340. doi: 10.1002/jmri.25970

20. Wang P, Nie P, Dang Y, Wang L, Zhu K, Wang H, Wang J, Liu R, Ren J, Feng J, Fan H, Yu J, Chen B. Synthesizing the First Phase of Dynamic Sequences of Breast MRI for Enhanced Lesion Identification. Front Oncol 2021;11:792516. doi: 10.3389/fonc.2021.792516

21. Kim E, Cho H-H, Kwon J, Oh Y-T, Ko ES, Park H. Tumor-Attentive Segmentation-Guided GAN for Synthesizing Breast Contrast-Enhanced MRI Without Contrast Agents. IEEE Journal of Translational Engineering in Health and Medicine 2022;11:32–43.

22. Muller-Franzes G, Huck L, Tayebi Arasteh S, Khader F, Han T, Schulz V, Dethlefsen E, Kather JN, Nebelung S, Nolte T, Kuhl C, Truhn D. Using Machine Learning to Reduce the Need for Contrast Agents in Breast MRI through Synthetic Images. Radiology 2023:222211. doi: 10.1148/radiol.222211

23. Sikka D, Zhu N, Liu C, Small S, Guo J. Predicting Gadolinium Contrast Enhancement for Structural Lesion Analysis using DeepContrast.

24. Liebert A, Schreiter H, Kapsner LA, Ohlmeyer S, Laun FB, Maier A, Uder M, Wenkel E, Bickelhaupt S. Virtual abbreviated contrast enhanced MRI for breast cancer diagnostics – initial experience. European Congress of Radiology. Vienna 2022.

25. Chung M, Calabrese E, Mongan J, Ray KM, Hayward JH, Kelil T, Sieberg R, Hylton N, Joe BN, Lee AY. Deep Learning to Simulate Contrast-enhanced Breast MRI of Invasive Breast Cancer. Radiology 2022:213199. doi: 10.1148/radiol.213199

26. Zhang T, Han L, D’Angelo A, Wang X, Gao Y, Lu C, Teuwen J, Beets-Tan R, Tan T, Mann R. Synthesis of Contrast-Enhanced Breast MRI Using T1-and Multi-b-Value DWI-Based Hierarchical Fusion Network with Attention Mechanism. Cham: Springer Nature Switzerland, 2023; p. 79–88.

27. Kapsner LA, Balbach EL, Folle L, Laun FB, Nagel AM, Liebert A, Emons J, Ohlmeyer S, Uder M, Wenkel E. Image quality assessment using deep learning in high b-value diffusion-weighted breast MRI. Scientific reports 2023;13(1):10549.

28. Kapsner LA, Ohlmeyer S, Folle L, Laun FB, Nagel AM, Liebert A, Schreiter H, Beckmann MW, Uder M, Wenkel E, Bickelhaupt S. Automated artifact detection in abbreviated dynamic contrast-enhanced (DCE) MRI-derived maximum intensity projections (MIPs) of the breast. Eur Radiol 2022. doi: 10.1007/s00330-022-08626-5

29. Bickelhaupt S, Tesdorff J, Laun FB, Kuder TA, Lederer W, Teiner S, Maier-Hein K, Daniel H, Stieber A, Delorme S. Independent value of image fusion in unenhanced breast MRI using diffusion-weighted and morphological T2-weighted images for lesion characterization in patients with recently detected BI-RADS 4/5 x-ray mammography findings. European radiology 2017;27:562–569.

30. Telegrafo M, Rella L, Stabile Ianora AA, Angelelli G, Moschetta M. Unenhanced breast MRI (STIR, T2-weighted TSE, DWIBS): An accurate and alternative strategy for detecting and differentiating breast lesions. Magn Reson Imaging 2015;33(8):951–955. doi: 10.1016/j.mri.2015.06.002

31. Fedorov A, Beichel R, Kalpathy-Cramer J, Finet J, Fillion-Robin JC, Pujol S, Bauer C, Jennings D, Fennessy F, Sonka M, Buatti J, Aylward S, Miller JV, Pieper S, Kikinis R. 3D Slicer as an image computing platform for the Quantitative Imaging Network. Magn Reson Imaging 2012;30(9):1323–1341. doi: 10.1016/j.mri.2012.05.001

32. Wang Z, Bovik AC, Sheikh HR, Simoncelli EP. Image quality assessment: from error visibility to structural similarity. IEEE transactions on image processing 2004;13(4):600–612.

33. Morley SK, Brito TV, Welling DT. Measures of model performance based on the log accuracy ratio. Space Weather 2018;16(1):69–88.

34. Baltzer P, Mann RM, Iima M, Sigmund EE, Clauser P, Gilbert FJ, Martincich L, Partridge SC, Patterson A, Pinker K, Thibault F, Camps-Herrero J, Le Bihan D. Diffusion-weighted imaging of the breast-a consensus and mission statement from the EUSOBI International Breast Diffusion-Weighted Imaging working group. Eur Radiol 2020;30(3):1436–1450. doi: 10.1007/s00330-019-06510-3

35. Ohlmeyer S, Laun FB, Bickelhaupt S, Palm T, Janka R, Weiland E, Uder M, Wenkel E. Ultra-High b-Value Diffusion-Weighted Imaging-Based Abbreviated Protocols for Breast Cancer Detection. Invest Radiol 2021;56(10):629–636. doi: 10.1097/rli.0000000000000784

36. Bickelhaupt S, Laun FB, Tesdorff J, Lederer W, Daniel H, Stieber A, Delorme S, Schlemmer HP. Fast and Noninvasive Characterization of Suspicious Lesions Detected at Breast Cancer X-Ray Screening: Capability of Diffusion-weighted MR Imaging with MIPs. Radiology 2016;278(3):689–697. doi: 10.1148/radiol.2015150425

37. Nogueira L, Brandão S, Matos E, Nunes RG, Loureiro J, Ramos I, Ferreira HA. Application of the diffusion kurtosis model for the study of breast lesions. European Radiology 2014;24(6):1197–1203. doi: 10.1007/s00330-014-3146-5

38. Jensen JH, Helpern JA. MRI quantification of non-Gaussian water diffusion by kurtosis analysis. NMR in Biomedicine 2010;23(7):698–710. doi: 10.1002/nbm.1518

39. Zimmermann F, Korzowski A, Breitling J, Meissner JE, Schuenke P, Loi L, Zaiss M, Bickelhaupt S, Schott S, Schlemmer HP, Paech D, Ladd ME, Bachert P, Goerke S. A novel normalization for amide proton transfer CEST MRI to correct for fat signal-induced artifacts: application to human breast cancer imaging. Magn Reson Med 2020;83(3):920–934. doi: 10.1002/mrm.27983

