## Supplement Material for "Impact of Non-Contrast Enhanced Imaging Input Sequences on the Generation of Virtual Contrast-Enhanced Breast MRI Scans using Neural Networks"

### MATERIALS AND METHODS

#### Literature Review

A search for relevant articles was performed in major databases, including PubMed, IEEE Xplore, ScienceDirect, and arXiv, as well as for conference abstracts in the International Society for Magnetic Resonance in Medicine annual meetings for the years 2020–2022 and the European Congress of Radiology 2022 using the following targeted search terms: “virtual contrast enhancement,” “synthetic contrast enhancement,” “breast MRI,” “neural networks,” and “generative adversarial network.” The selection criteria were based on the relevance of the studies to our research question, with a focus on original research articles that reported the development and evaluation of neural network-based vCE imaging methods for breast MRI. Based on this investigation, seven different publications and conference abstracts were identified.

Following the identification of the relevant publications, we extracted and compared the details of the employed MRI sequences (e.g., T1-weighted, T2-weighted, and diffusion-weighted imaging), neural network architectures (e.g., encoder–decoder convolutional neural networks or GANs), number of patients (with distributions into training, validation, and test datasets), and quantitative and qualitative evaluation methods.

#### Neural Network Architecture and Training

A graphical depiction of the network is shown in Appendix Figure 1. The networks were trained on a dedicated workstation (Linux Ubuntu 20.04, Intel Xeon E5-2698, 2.20 GHz, 48 Cores, 256 GB RAM) using a single NVIDIA v100 GPU card with 32 GB RAM. During training, batches of 35 slices were used, and, in each batch, it was ensured that random slices from different examinations were present. The network was trained for 35 epochs without early stopping. The learning rate during the training was set to $10^{-3}$. The neural network model from the epoch with the lowest value of the loss function over the validation set was used as the final model to generate the image series of the test set.

An adjusted loss function from the work of Chen et al. (1) was used during training. As our data lacked the segmentation of lesions, a combination of the structural similarity index and L1 norm was noted, as presented in the equation below.

$$L(y,\hat{y}) = \left(1 - [l(y,\hat{y})]^{\alpha} \cdot [c(y,\hat{y})]^{\beta} \cdot [s(y,\hat{y})]^{\gamma}\right) + \sum_{i=0}^{N} |y_i - \hat{y}_i|$$

The network was implemented using Python (version 3.8.10) via the PyTorch (version 1.9.0) framework.

#### Rationale for the Exclusion of ADC and Simulated Low-Dose Images

We did not use ADC maps as input data, as they are prone to errors introduced during ADC calculation. We believe that neural networks themselves can derive the same information from raw DWI data without the need for ADC maps. Additionally, as suggested by Muller-Franzes et al. (2), the simulated low-dose images were not used, as these images are not based solely on native input sequences. The simulated low-dose images require a preprocessing of a contrast-enhanced MRI acquisition, which deviates from our goal of investigating the influence of only native MRI input sequences.

#### Segmentation of Target Findings

The target findings were segmented by a medical student with 2 years of experience in breast MRI research under the supervision of a board-certified radiologist with >10 years of experience using the open-source 3D Slicer Software's [version 4.11, Fedorov et al. (3)] built-in region draw function. The target findings were selected in consensus based on the clinical routine report and then segmented manually using the original post-contrast images as matrix and the full multiparametric information of the MRI examinations (e.g., in case of non-enhancing lesions such as cysts, in which, however, contours of the cysts can be delineated carefully when shifting co-registered images in between T1-weighted post-contrast subtraction and T2-weighted sequences). The segmentations were performed along the inner border of the target finding, suing the slice depicting the target finding most centrally. In case of multiple target findings, only the largest target finding within the breast volume was segmented. The size of the target finding was measured as the length of the longest axis in the segmented target finding. For the analysis of the target finding regarding the performance metrics, a bounding box around the segmentation was drawn using the skimage Python package. The metrics were then calculated only inside the bounding box.

#### Quantitative Metrics Choice and Calculation

The literature review (Table 1) showed that the two most commonly used similarity metrics were the SSIM and PSNR. The SSIM was calculated in accordance with the work of Wang et al. (4). The error metrics selected were the normalized root mean square error (NRMSE) and median symmetrical accuracy (MEDSYMAC). We used the MEDSYMAC instead of the SMAPE or MAPE, as subtraction images commonly consist of many substantially low values; the SMAPE and MAPE could then potentially overestimate the error, as they are prone to values close to 0.

#### Inter-Reader Agreement Evaluation

Inter-reader agreement between the Likert scale-assessed variables was evaluated using Kendall's coefficient of concordance.

### RESULTS

#### Inter-Reader Agreement Evaluation

Kendall's coefficient of concordance showed the highest agreement between the three readers for the lesion enhancement score (mean=0.94±0.01), followed by the satisfaction with image contrast and SNR (mean=0.88±0.04). The image quality and sharpness showed lower but still good agreement between the readers (mean=0.82±0.04 and 0.80±0.04, respectively).

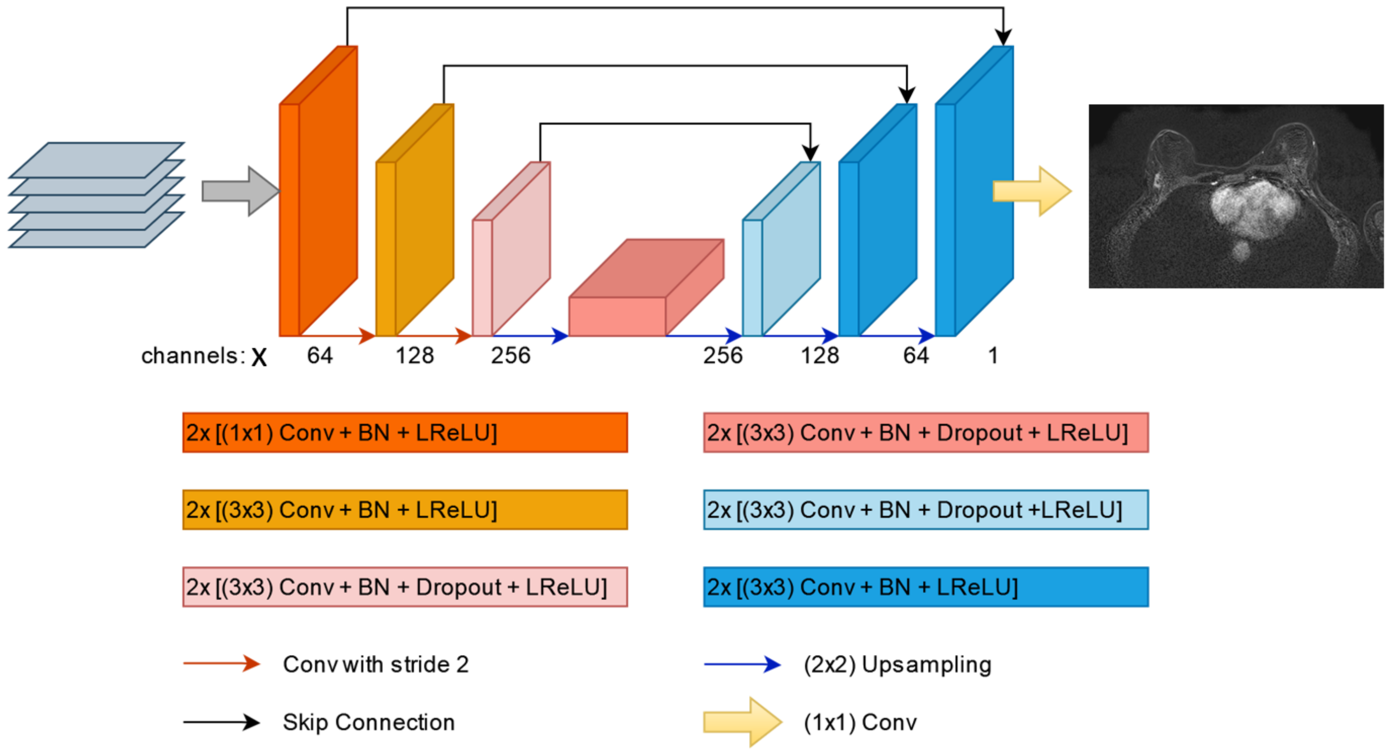


**Supplemental Figure 1:** A 2D U-net architecture consisted of three encoder and three decoder stages with a bottleneck layer between the deepest encoder and decoder stages followed by an output stage. The number of the networks input channels (x) depends on the specific input sequence combination.
Each encoder and decoder stage consisted of two convolutional layers with a convolutional kernel size of 3, followed by batch normalization and leaky rectified linear unit (LReLU) activation function. The encoder and decoder stages connected to the bottleneck layer had additional dropout layers between the batch normalization layer and the activation function layer. The down- and up-sampling of the spatial size and feature maps was performed via a 2×2 convolution and a transposed 2×2 convolution with a stride of 2, respectively. The initial encoder stage, was set to generate 64 features, resulting in a maximal feature size of 512. The output layer consisted of 1×1 convolution layer, reducing the number of output channels to 1, and was followed by a tanh function to map the output results to [−1,1].

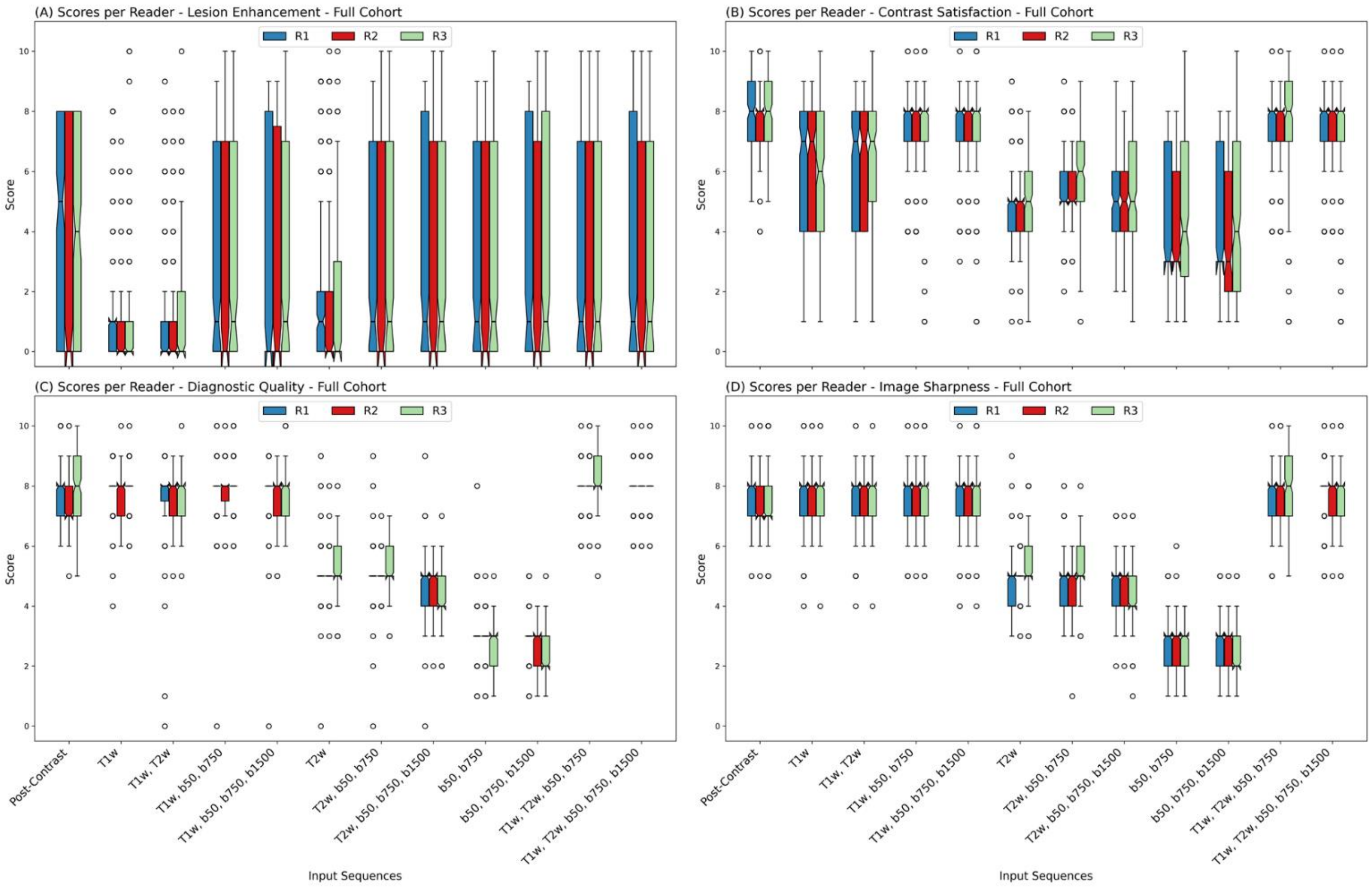


**Supplement Figure 2:** Individual qualitative evaluation scores among the three readers (R1, R2, and R3) for the full patient cohort. A significant drop in the ability to enhance lesions/NMEs could be observed for all three readers when no DWI acquisition was present in the input sequence combination. A significant drop in the diagnostic quality and image sharpness score could be noted when no T1w acquisition was present in the input sequence combination for all three readers. A significant drop in the contrast satisfaction could be observed when either the T1w image acquisition or DWI acquisition was missing in the input sequence combination. T1w=T1-weighted, b50=DWI acquisition with a b-value of 50 s/mm$^2$, b750=DWI acquisition with a b-value of 750 s/mm$^2$, b1500=DWI acquisition with a b-value of 1500 s/mm$^2$, NME=non-mass-enhancement

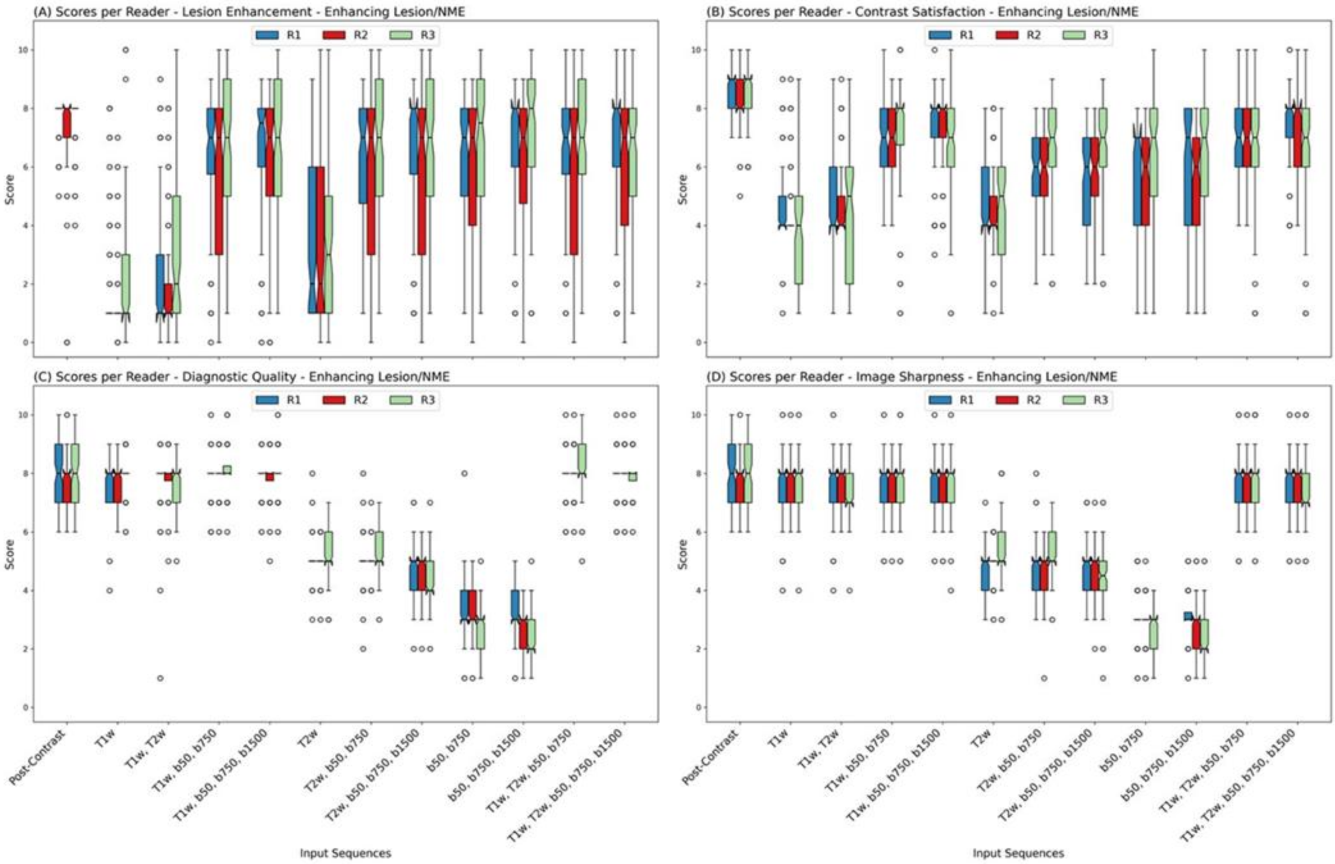


**Supplement Figure 3:** Individual qualitative evaluation scores among the three readers (R1, R2, and R3) for only the cases with enhancing lesions/NMEs in the original post-contrast subtraction image after GBCA injection. A significant drop in the ability to enhance lesions/NMEs could be observed for all three readers when no DWI acquisition was present in the input sequence combination for all three readers when compared with the original post-contrast image. A significant drop in the diagnostic quality and image sharpness score could be noted when no T1w image acquisition was present in the input sequence combination for all three readers. For all three readers, a significant drop in the contrast satisfaction could be observed when either the T1w image acquisition or DWI acquisition was missing in the input sequence combination. Further, for all three readers, an overall lower score of contrast satisfaction could be noted even for input combinations that included both T1w image acquisition and all three DWI acquisitions. T1w=T1-weighted, b50=DWI acquisition with a b-value of 50 s/mm$^2$, b750=DWI acquisition with a b-value of 750 s/mm$^2$, b1500=DWI acquisition with a b-value of 1500 s/mm$^2$, NME=non-mass-enhancement

**Supplement Table 1: Quantitative Performance of the Different Network Input Combinations**

| Input Combination | Tissue | SSIM (↑) | PSNR [dB] (↑) | NRMSE [%] (↓) | MEDSYMAC [%] (↓) |
|---|---|---|---|---|---|
| T1w | BV | 86.91±2.58 | 24.18±1.82 | 8.91±1.20 | 2.08±0.95 |
| | TF* | 50.25±22.84 | 16.77±6.10 | 20.05±6.63 | 13.92±5.19 |
| T1w+T2w | BV | 87.06±2.57 | 24.33±1.79 | 8.77±1.17 | 2.02±0.92 |
| | TF* | 51.94±22.80 | 17.15±6.17 | 20.41±7.49 | 14.12±6.28 |
| T1w, b50/b750 | BV | 86.98±2.60 | 24.25±1.75 | 8.86±1.18 | 1.78±0.85 |
| | TF* | 60.63±20.71 | 18.51±5.25 | 19.82±7.46 | 11.88±5.58 |
| T1w, b50/b750/b1500 | BV | 87.00±2.55 | 24.26±1.83 | 8.79±1.20 | 2.09±0.92 |
| | TF* | 63.08±20.16 | 19.02±5.24 | 19.06±7.83 | 11.01±5.07 |
| T2w | BV | 78.18±5.13 | 22.82±1.70 | 10.44±1.32 | 3.38±1.26 |
| | TF* | 44.29±21.58 | 16.34±5.28 | 25.43±7.89 | 15.69±5.72 |
| T2w, b50/b750 | BV | 78.41±5.07 | 23.00±1.75 | 10.28±1.37 | 3.36±1.28 |
| | TF* | 52.02±21.43 | 17.54±4.85 | 23.37±8.06 | 13.69±5.63 |
| T2w, b50/b750/b1500 | BV | 78.44±5.09 | 22.92±1.75 | 10.36±1.39 | 3.07±1.23 |
| | TF* | 52.90±21.21 | 17.58±4.97 | 23.36±8.10 | 13.82±5.62 |
| b50/b750 | BV | 77.38±4.92 | 22.50±1.68 | 10.90±1.32 | 3.85±1.35 |
| | TF* | 50.16±22.74 | 16.59±4.81 | 26.21±8.32 | 15.51±5.59 |
| b50/b750/b1500 | BV | 77.41±4.93 | 22.51±1.73 | 10.89±1.36 | 3.89±1.33 |
| | TF* | 51.17±22.79 | 16.45±4.90 | 26.01±9.84 | 14.98±5.67 |
| T1w, T2w, b50/b750 | BV | 87.12±2.58 | 24.38±1.80 | 8.73±1.18 | 1.94±0.87 |
| | TF* | 60.24±20.46 | 18.75±5.55 | 19.94±8.39 | 11.70±5.60 |
| T1w, T2w, b50/b750/b1500 | BV | 87.10±2.55 | 24.36±1.78 | 8.74±1.17 | 1.91±0.87 |
| | TF* | 61.11±20.24 | 18.16±5.57 | 19.67±8.21 | 11.46±5.17 |

T1w=T1-weighted, T2w=T2-weighted, b50=diffusion-weighted imaging with a b-value of 50 s/mm$^2$, b750=diffusion-weighted imaging with a b-value of 750 s/mm$^2$, b1500=diffusion-weighted imaging with a b-value of 1500 s/mm$^2$, BV=breast volume, TF – Target findings, SSIM=structural similarity index, PSNR=peak signal-to-noise ratio, dB=decibel, NRMSE=normalized root mean square error, MEDSYMAC=median symmetrical accuracy

*Target findings refer to the findings within the examination that could be both benign or malignant and non-mass or mass enhancement as well as findings not enhancing (e.g., cysts) but morphologically delineated from healthy fibroglandular tissue.

### REFERENCES


1. Chen C, Raymond C, Speier B, Jin X, Cloughesy TF, Enzmann D, Ellingson BM, Arnold CW. Synthesizing MR Image Contrast Enhancement Using 3D High-resolution ConvNets. arXiv preprint arXiv:210401592 2021.
2. Muller-Franzes G, Huck L, Tayebi Arasteh S, Khader F, Han T, Schulz V, Dethlefsen E, Kather JN, Nebelung S, Nolte T, Kuhl C, Truhn D. Using Machine Learning to Reduce the Need for Contrast Agents in Breast MRI through Synthetic Images. Radiology 2023:222211. doi: 10.1148/radiol.222211
3. Fedorov A, Beichel R, Kalpathy-Cramer J, Finet J, Fillion-Robin JC, Pujol S, Bauer C, Jennings D, Fennessy F, Sonka M, Buatti J, Aylward S, Miller JV, Pieper S, Kikinis R. 3D Slicer as an image computing platform for the Quantitative Imaging Network. Magn Reson Imaging 2012;30(9):1323-1341. doi: 10.1016/j.mri.2012.05.001
4. Wang Z, Bovik AC, Sheikh HR, Simoncelli EP. Image quality assessment: from error visibility to structural similarity. IEEE transactions on image processing 2004;13(4):600-612.
